# The angiotensin type 2 receptor agonist C21 restores respiratory function in COVID19 - a double-blind, randomized, placebo-controlled Phase 2 trial

**DOI:** 10.1101/2021.01.26.21250511

**Authors:** Göran Tornling, Rohit Batta, Joanna Porter, Thomas Bengtsson, Kartikeya Parmar, Reema Kashiva, Anders Hallberg, Anne Katrine Cohrt, Kate Westergaard, Carl-Johan Dalsgaard, Johan Raud

**Author notes:** **Correspondence:** Carl-Johan Dalsgaard, Vicore Pharma AB, Kronhusgatan 11, SE-411 05 Gothenburg, Sweden.

## Abstract

**Background:** Although several therapies have been evaluated for treatment of COVID-19, the morbidity and mortality in COVID-19 are still significant, and the need for safe and effective drugs remains high even after launch of vaccine programs.

**Methods:** We conducted a double-blind, randomized, placebo-controlled trial with the novel oral angiotensin II type 2 receptor agonist C21 in hospitalized COVID-19 patients with C-reactive protein 50-150 mg/L but not needing mechanical ventilation. Patients were randomly assigned to oral C21 (100 mg twice daily) or placebo for 7 days in addition to standard of care, including glucocorticoids and remdesivir.

**Results:** 106 patients underwent randomization (51 in the C21 group and 55 in the placebo group). At day 14 after start of treatment, the proportion of patients still requiring supplemental oxygen was significantly reduced by 90% in the C21 group compared to the placebo group (p=0.003). Moreover, fewer patients required mechanical ventilation (one C21 patient and four placebo patients), and C21 was associated with a numerical reduction in the mortality rate (one and three deaths in the C21 and placebo group, respectively). Treatment with C21 was safe and well tolerated.

**Conclusions:** As studied in hospitalized COVID-19 patients, C21 on top of standard of care led to a clinically beneficial improvement in respiratory function compared to placebo, paving the way for a pivotal randomised controlled trial.

This study is registered at ClinicalTrials.gov with identifier NCT04452435.

## INTRODUCTION

In January 2020, SARS-CoV-2 was identified as the causative agent of an outbreak of the new viral pneumonia disease COVID-19, with the first cases reported in December 2019 (1-3). The disease expanded rapidly, and by 12 March 2020, COVID-19 was classified as a pandemic by the World Health Organization (WHO). As of 25 January 2021, the WHO has reported almost 100 million confirmed cases of COVID-19 and more than 2 million deaths globally (4).

While most COVID-19 cases result in mild disease only, a substantial proportion of patients develop severe respiratory illness resulting in impaired gas exchange, hypoxia, need of supplementary oxygen and, in the most severe cases, mechanical ventilation (3, 5, 6).

To date, only two drugs, remdesivir and dexamethasone, have shown some impact on disease remission in large controlled trials in hospitalized COVID-19 patients and are now part of current standard of care (7, 8), Despite the benefits of these drugs, the morbidity and mortality in COVID-19 are still significant, particularly in the moderately severe disease where it could be argued that no therapies have shown a consistent meaningful benefit (9). The need for safe, effective and convenient COVID-19 drugs is likely to remain even after the launch of vaccine programs.

The SARS-CoV-2 virus is known to bind to and enter target cells through angiotensin converting enzyme 2 (ACE2), an integral component of the renin-angiotensin system (RAS) (3). Within the RAS, the angiotensin II (AngII) type 1 receptor (AT1R) normally mediates the classical actions of AngII, including constriction of blood vessels, sodium retention and cell growth, while abnormal AT1R activation contributes to the pathogenesis of certain cardiovascular, renal and pulmonary diseases (10-12). Conversely, activation of the angiotensin II type 2 receptor (AT2R) causes dilatation of blood vessels and inhibition of inflammation and fibrosis and is considered to be counter-regulatory to the negative effects of AT1R activation (12, 13). Natural ligands/agonists of AT2R such as Ang 1-7 and Ang 1-9 are products of ACE2 cleaving AngII (14). As the binding of SARS-CoV-2 virus to ACE2 is understood to downregulate and inactivate ACE2, we speculated that such SARS-CoV-2-induced down-regulation may result in the RAS being thrown out of balance. That this ACE2 deactivation may lead to overstimulation of the AT1R and understimulation of the AT2R in COVID-19 has been discussed in a recent review article by Steckelings and Sumners (15).

The AT2R is mainly expressed in embryonic tissue and, under normal conditions, only at low levels in most tissues in healthy adults (16, 17). However, it has recently been described that the AT2R is highly expressed in alveolar type 2 (AT2) progenitor cells in the adult human lung (18) and can be upregulated during repair and regeneration (19). Moreover, the AT2 cells are the primary site of SARS-CoV-2 replication in the distal airways (20) which is likely to contribute to the alveolar dysfunction and impaired gas exchange caused by the virus.

Compound 21 (C21) is a first-in-class, low molecular weight, orally available, specific, high-affinity AT2R agonist (21) currently in clinical development in idiopathic pulmonary fibrosis (22). In recent phase I safety and pharmacokinetics studies, 100 mg C21 twice daily was found to be safe and well tolerated (see Clinical Trial Protocol available with the full text of this article).

Based on the established safety and tolerability of C21 in healthy subjects and the unprecedented imperative for new and effective treatments of patients with severe COVID-19, we investigated whether C21 could have beneficial effects in this disease. To our knowledge, this is also the first study of an AT2R agonist in any human disease.

## METHODS

### Design

Enrolment for this trial named ATTRACT (Angiotensin II Type Two Receptor Agonist COVID-19 Trial) was performed between 21 July and 29 September 2020 at 8 trial sites in India. The trial enrolled 19 to 69 years old patients who were hospitalized with coronavirus (SARS-CoV)-2 infection confirmed by polymerase chain reaction (PCR) test <4 days before enrolment and with signs of an acute respiratory infection but not requiring invasive or non-invasive mechanical ventilation on the day of randomization. To be eligible for inclusion in the trial, the patient should have a C-reactive protein (CRP) value of >50 and <150 mg/L. Concomitant medication and supportive care as per standard of care at the trial site was permitted, although patients should not have received any previous experimental treatment for COVID-19. Informed consent was obtained from each patient prior to any trial-related procedure. Full inclusion and exclusion criteria are presented in the Supplement.

The patients were randomly assigned in a 1:1 ratio to receive either C21 or placebo in blocks of four, with randomization stratified by trial site. Patients, care providers and those assessing outcomes were kept blinded until data base lock had been performed for the complete trial. Twice daily doses of 100 mg C21 or matching placebo were administered orally for 7 days. A final visit (at the hospital or by phone call) was performed 7-10 days after the last dose of the trial drug. The trial drug was withdrawn if the patient needed mechanical invasive or non-invasive ventilation or was discharged early from the hospital. Patients were withdrawn from the trial if the Investigator judged it necessary due to medical reasons (e.g. adverse events), if lost to follow-up or at the patient’s own decision. Full withdrawal criteria are presented in the Supplement.

### Procedures and Outcomes

During the hospitalization, patients were assessed daily by physical examination, vital signs, need for supplementary oxygen, body temperature, urine analysis and blood tests for CRP and other biomarkers (IL-6, IL-10, TNF, CA125 and ferritin).

The primary endpoint of the trial, change in CRP from baseline to end-of-treatment, was based on early data in glucocorticoid naïve patients demonstrating that CRP predicts severe outcomes. Secondary endpoints included need for supplementary oxygen, need for mechanical invasive or non-invasive ventilation, other biomarkers and adverse events. Full description of endpoints is available in the Supplement.

### Statistical analyses

The original sample size calculation on the primary endpoint indicated that 75 patients per group were needed to achieve 80% power to detect a difference in reduction of CRP in C21 treated patients compared to placebo using a two-sided t-test at 10% significance level. Due to recruitment challenges, the recruitment was halted after 106 patients were randomized.

For biomarkers, the mean of the last two assessments in the treatment period was used for analysis. Data was compared between treatments using ANCOVA with treatment as factor and baseline as covariate. Data were logged prior to analysis. Patients not in need of supplementary oxygen or not in need of mechanical ventilation were compared using logistic regression. Proportion of days on supplemental oxygen was compared using the Wilcoxon rank sum test. Patients with no baseline or no post-treatment data were excluded from analysis of biomarkers. Patients in ventilator care/deaths were considered in oxygen need from time of withdrawal. For other withdrawals, the last value assessed was carried forward. The Statistical Analysis Plan is available with the full text of this article.

In *post hoc* analyses, data on oxygen supplementation up to 14 days after start of treatment were analyzed, and sub-analyses based on disease severity at randomisation as assessed by need for supplementary oxygen were performed by Chi-square test.

### Ethical and regulatory authority approval

The protocol, patient information, patient consent form and other documents, as required, were approved by properly constituted IECs and by the national regulatory authorities. All patients gave written informed consent. The trial was registered at ClinicalTrials.gov, number NCT04452435.

## RESULTS

### Patients

Of the 206 patients who were assessed for eligibility, 106 underwent randomization; 51 were assigned to the C21 group and 55 to the placebo group (Figure 1). Six patients had C21 treatment discontinued before completion of all doses due to need for mechanical ventilation (1 patient), withdrawal of consent (1) or discharge from hospital (4). Twelve patients had placebo treatment discontinued before completion of all doses and one during the follow-up period due to need for mechanical ventilation (4 patients), withdrawal of consent (4) or discharge from hospital (5).

**Figure 1.**
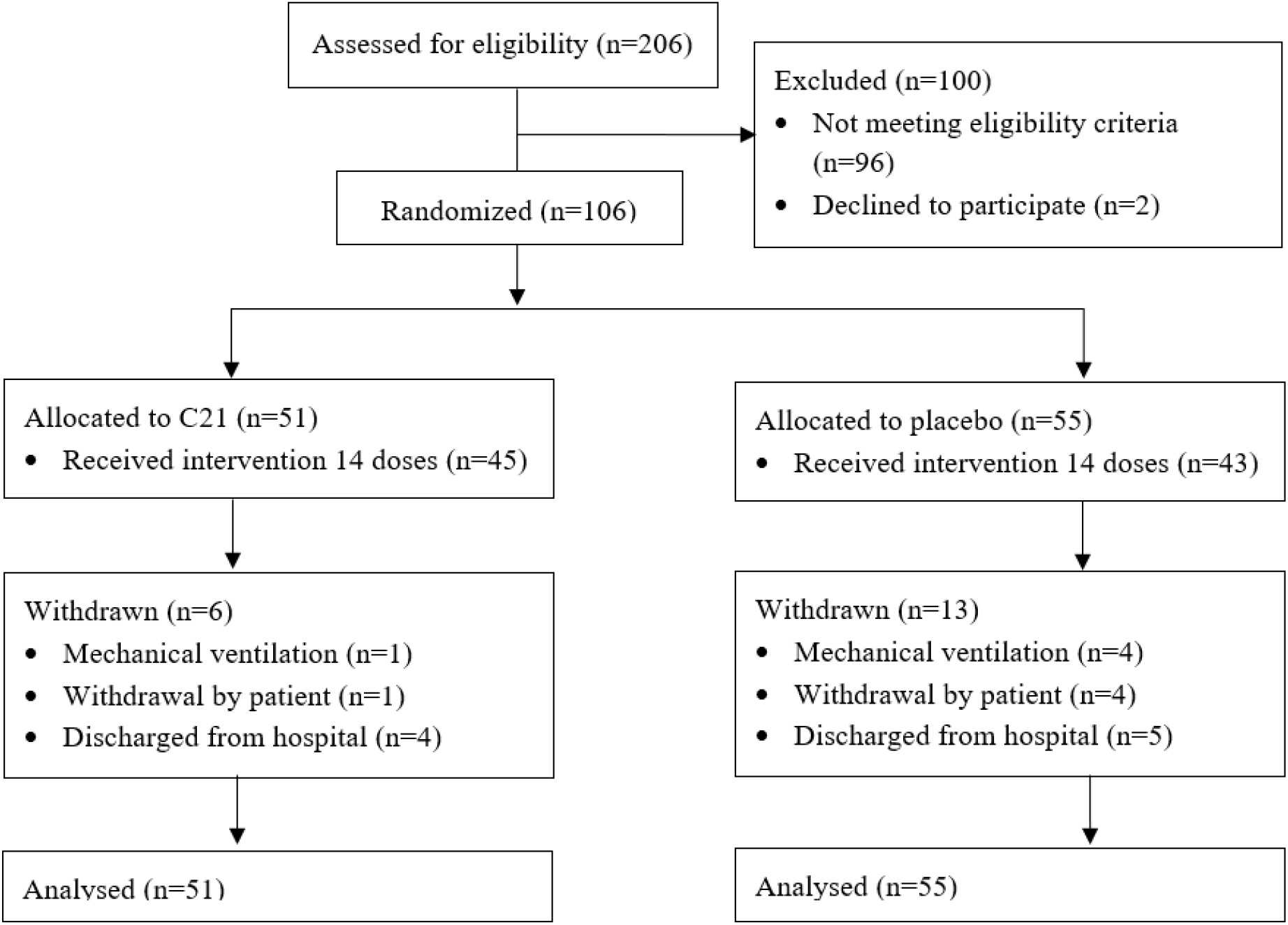
Enrollment and Randomization.

As shown in Table 1, the treatment groups were well balanced regarding age (mean 52.6 years) and gender (75.5% males). Most patients had one or more coexisting medical condition. Hypertension and overweight/obesity were more common in the C21 group compared with the placebo group, but there was no difference in the presence of diabetes mellitus. Supplementary oxygen supply at baseline was needed in 57.5% of the patients with no major difference between the treatment groups.

**Table 1.** Demographic and Clinical Characteristics at Baseline.

| Characteristic | C21<br>(N=51) | Placebo<br>(N=55) | All<br>(N=106) |
| --- | --- | --- | --- |
| Age; mean (SD) | 54.3 (9.13) | 51.1 (11.18) | 52.6 (10.30) |
| Male sex; N (%) | 38 (74.5) | 42 (76.4) | 80 (75.5) |
| Body mass index (kg/m<sup>2</sup>); mean (SD) | 25.4 (4.03) | 25.1 (3.43) | 25.2 (3.71) |
| Overweight (30>BMI≥25.0); N (%) | 22 (43.1) | 18 (32.7) | 40 (37.7) |
| Obese (BMI≥30.0); N (%) | 4 (7.8) | 6 (10.9) | 10 (9.4) |
| Overweight or obese (BMI≥25.0); N (%) | 26 (51.0) | 24 (43.6) | 50 (47.2) |
| Coexisting conditions; N (%) |  |  |  |
| Diabetes mellitus | 17 (33.3) | 19 (34.5) | 36 (34.0) |
| Hypertension | 18 (35.3) | 14 (25.5) | 32 (30.2) |
| Supplemental oxygen use at Baseline; N (%) | 29 (56.9) | 32 (58.2) | 61 (57.5) |

Concomitant medication according to local standard of care was permitted in the trial. A vast majority of the patients received glucocorticoids, and most patients received antibacterial and antiviral compounds (remdesivir in 67.0%), with no major differences between the treatment groups (Table 2).

**Table 2.**
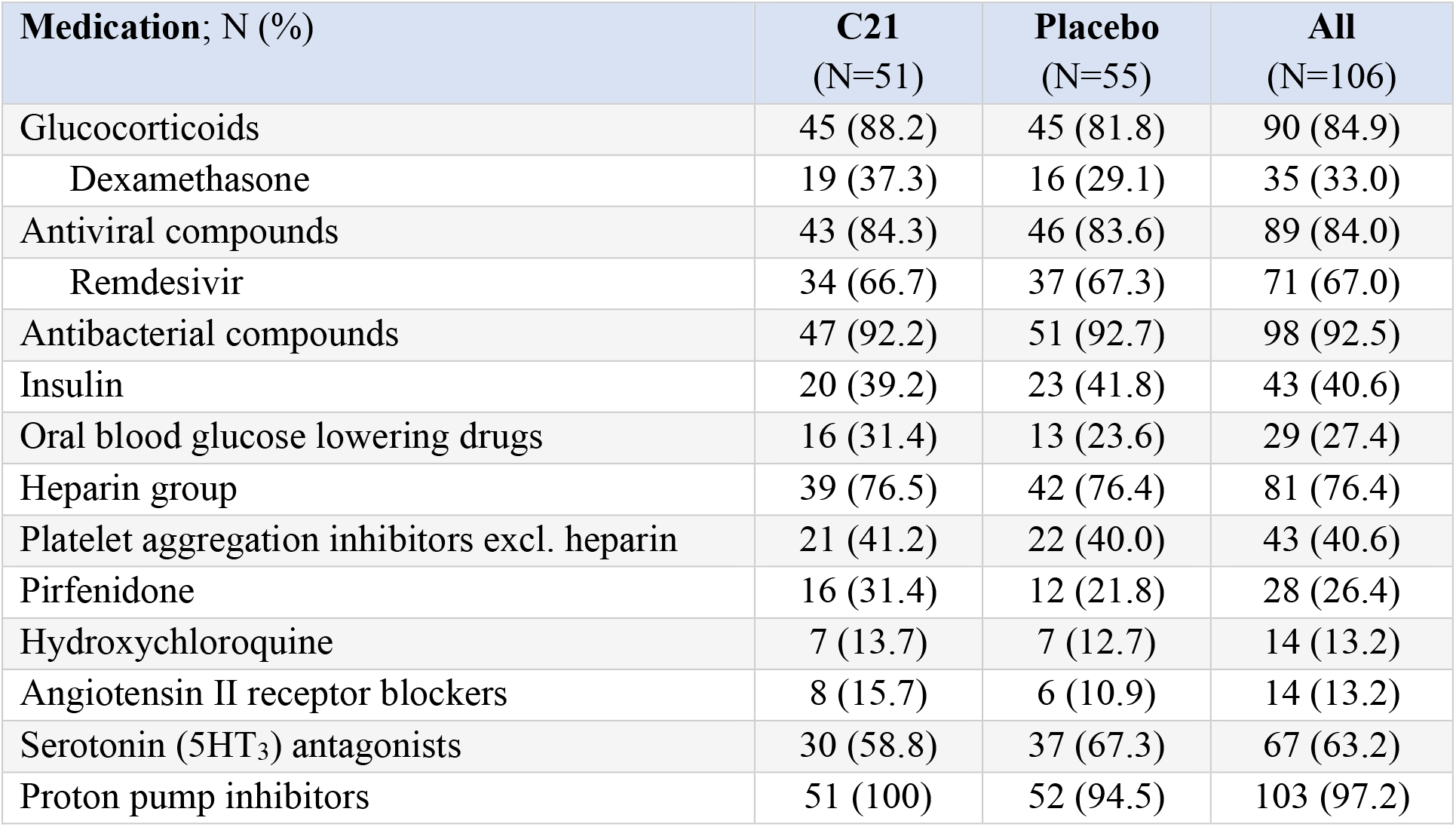
Concomitant medication.

| **Medication; N (%)** | **C21**<br>**(N=51)** | **Placebo**<br>**(N=55)** | **All**<br>**(N=106)** |
| --- | --- | --- | --- |
| Glucocorticoids | 45 (88.2) | 45 (81.8) | 90 (84.9) |
| Dexamethasone | 19 (37.3) | 16 (29.1) | 35 (33.0) |
| Antiviral compounds | 43 (84.3) | 46 (83.6) | 89 (84.0) |
| Remdesivir | 34 (66.7) | 37 (67.3) | 71 (67.0) |
| Antibacterial compounds | 47 (92.2) | 51 (92.7) | 98 (92.5) |
| Insulin | 20 (39.2) | 23 (41.8) | 43 (40.6) |
| Oral blood glucose lowering drugs | 16 (31.4) | 13 (23.6) | 29 (27.4) |
| Heparin group | 39 (76.5) | 42 (76.4) | 81 (76.4) |
| Platelet aggregation inhibitors excl. heparin | 21 (41.2) | 22 (40.0) | 43 (40.6) |
| Pirfenidone | 16 (31.4) | 12 (21.8) | 28 (26.4) |
| Hydroxychloroquine | 7 (13.7) | 7 (12.7) | 14 (13.2) |
| Angiotensin II receptor blockers | 8 (15.7) | 6 (10.9) | 14 (13.2) |
| Serotonin (5HT<sub>3</sub>) antagonists | 30 (58.8) | 37 (67.3) | 67 (63.2) |
| Proton pump inhibitors | 51 (100) | 52 (94.5) | 103 (97.2) |

### Clinical outcomes

Extended need for oxygen therapy was more frequent in the placebo group than in the C21 group (Figure 2 and Table E2). At Day 14, only one patient in the C21 group, compared to 11 patients in the placebo group, needed supplementary oxygen (p=0.003). This difference was already apparent at the end of the treatment period, when 27.5% of patients in the C21 group and 45.5% in the placebo group were in need for supplemental oxygen (p=0.055).

**Figure 2.**
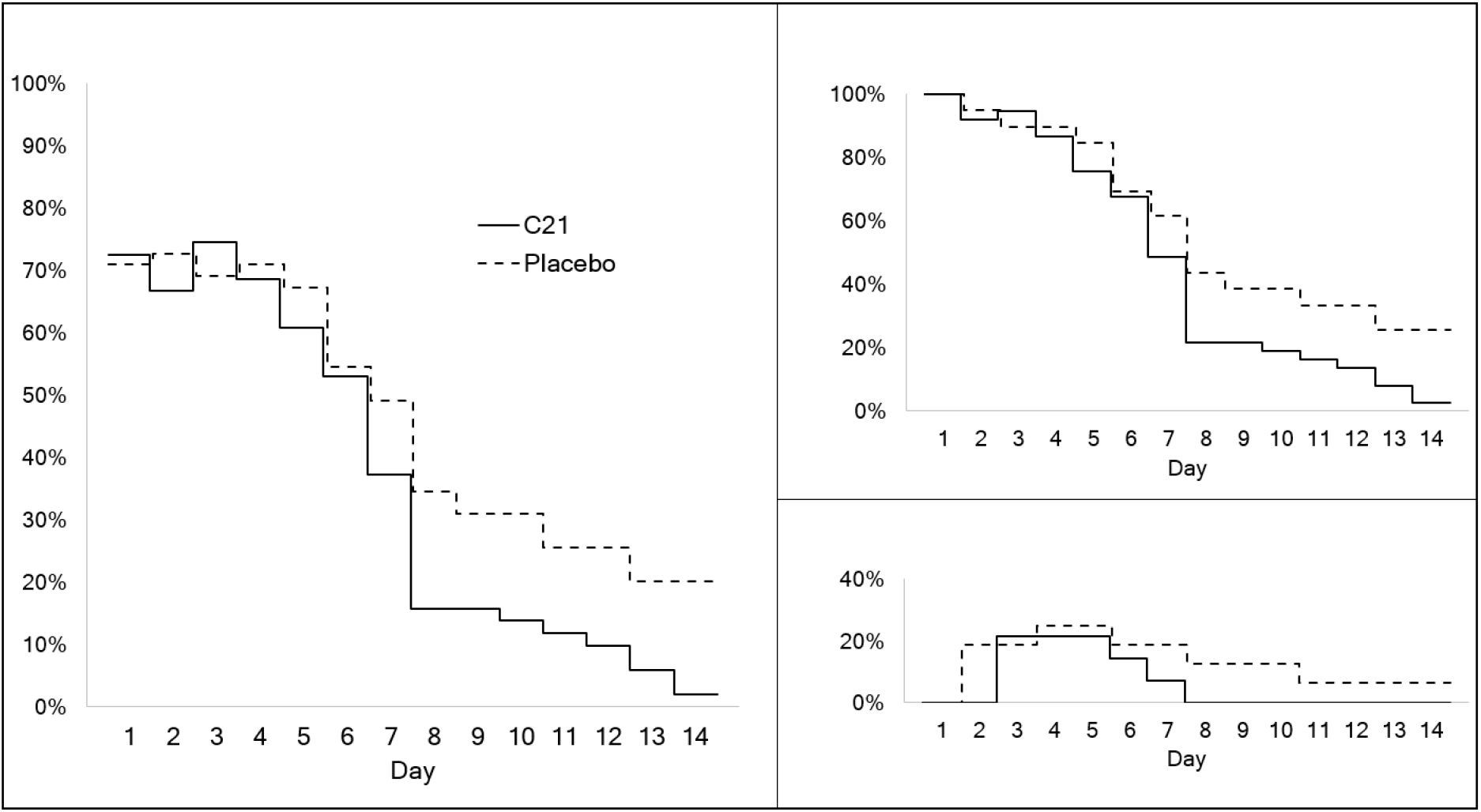
Need for supplemental oxygen therapy during the treatment and follow-up periods. Daily estimates of the proportion of patients in need for supplemental oxygen therapy. The left panel shows the total intention to treat population (C21 N=51, placebo N=55). The upper right panel shows patients in need for supplemental oxygen on the day of randomization (C21 N=37, placebo N=39), and the lower right panel shows patients not in need for oxygen on the day of randomization (C21 N=14, placebo N=16).

There were 4 deaths in the trial, one in the C21 group and 3 in the placebo group. All deaths occurred in patients with progressive respiratory insufficiency and need for mechanical ventilation. One more patient in the placebo group deteriorated and developed need for mechanical ventilation (4 events of mechanical ventilation in the placebo group vs 1 event in the C21 group). There were 64 treatment-emergent adverse events reported by 60.8% of the patients in the C21group and 90 events reported by 67.3% of the patients in the placebo group (Table E3). Most events were mild, and no event in either group was classified as related to trial treatment by the investigators. The most frequent adverse event was hyperglycemia, which occurred more frequently in the C21 group than in the placebo group. The level of hyperglycemia was modest, and generally not associated with glucosuria which was more frequently reported in the placebo group (Table E3). Further, there was no increased proportion of patients in the C21 group needing insulin compared to the placebo group (Table 2).

### Biomarkers

In both the C21 and the placebo group, CRP decreased rapidly between screening and randomization/baseline, and from randomization to end of treatment there was a further continued reduction with 81% and 78% in the C21 and placebo group, respectively (p=0.489, Table E1). Thus, there was little room for further CRP reduction by C21 and the primary endpoint of a change from baseline in CRP was not met, most likely due to the concomitant high use of glucocorticoids in both arms following the release of the RECOVERY trial (8). Nevertheless, in the more severely sick patients who needed oxygen therapy at baseline (57.5%), the difference was more pronounced with a statistically significant (at the predefined 10% level) CRP reduction of 84% in the C21 group and 72 % in the placebo group at the end of the treatment period (p=0.088, Table E1). There was no difference in effect on other biomarkers (IL-6, IL-10, TNF, CA125 and ferritin) between C21 and placebo (Table E1).

## DISCUSSION

This double-blind, randomized, placebo-controlled trial demonstrated that the AT2R agonist C21 was beneficial in the treatment of COVID-19, with patients receiving C21 having a significantly lower risk for extended need of supplementary oxygen therapy. The data also suggest that treatment with C21 may have prevented the progression to more severe respiratory disease, as shown by the lower proportion of need for mechanical ventilation or death due to respiratory failure. Importantly, C21 was given on top of standard of care, with a vast majority of patients receiving glucocorticoids and two thirds receiving remdesivir. The trial, which is the first study of an AT2R agonist in any human disease, also demonstrated that C21 was safe and well tolerated in patients with COVID-19.

The reduced need for extended oxygen supplementation indicates that treatment with C21 improves gas exchange at the alveolar level. The AT2 progenitor cells, apparently the only cells in the lungs that express the AT2R (18), are the primary site for viral replication in the distal airways (20), and it is suggested that C21 restores lung function by acting directly on these cells.

The effect of C21 did not seem to be related to an overt anti-inflammatory action since it was effective on top of glucocorticoid treatment and without further suppressing the proinflammatory cytokines IL-6 and TNF. CRP was initially selected as the primary endpoint because CRP has been reported to predict severe disease (23, 24) and mortality (6) in COVID-19. However, there was a rapid and profound decrease in CRP in both the C21 and the placebo group, most likely because the patients were treated with glucocorticoids which became part of standard of care at the time of trial initiation. Nevertheless, in the patients with more pronounced respiratory distress with need of supplementary oxygen at baseline, there was a small but significantly greater reduction of CRP in the C21 than in the placebo group.

Viral pneumonias can result in long-standing complications, including pulmonary fibrosis (25, 26). In a follow-up investigation of 133 patients 100 days after the diagnosis of COVID-19, lung function impairment was seen in 36% and pathological CT findings in 63% (27). Severity and duration of the initial phase of the disease seem to be important, since days on oxygen supplementation during the acute phase of COVID-19 was identified as a risk indicator for decreased diffusion capacity and increased total CT score at 12 weeks after the initial infection (28). COVID-19 shares characteristics with the severe acute respiratory syndrome (SARS), and the Middle East respiratory syndrome-coronavirus (MERS-CoV), and long-term follow-up studies in both these diseases have demonstrated a high frequency of pulmonary fibrosis (29, 30). However, the experience from the SARS and MERS cannot be directly transferred to COVID-19 due to the differences in demography as COVID-19 affects an older population which may lead to a higher frequency of long-term sequelae. Anti-fibrotic therapies, primarily nintedanib and pirfenidone, have been suggested for the prevention of fibrosis after SARS-CoV-2 infection (31). C21 has shown efficacy in pre-clinical studies of pulmonary fibrosis (32, 33), which suggests that it may have a broader use in longer term treatment of COVID-19 pneumonia, its sequelae and/or so-called ‘long COVID’.

In conclusion, C21 on top of standard of care, including glucocorticoids and remdesivir, significantly improved respiratory function reflected by a reduced need for supplemental oxygen in hospitalized COVID-19 patients. A further pivotal study of C21 in this disease is warranted.

## Supporting information

Clinical Trial Protocol

Statistical Analysis Plan

## Data Availability

Data will be made available to researchers who provide a scientifically sound proposal. Proposals should be directed to

## Acknowledgments

We thank all the participants in this clinical trial; the ATTRACT trial team, including all the investigators at the clinical sites; Mimi Flensburg (previous Vicore Pharma employee) for the instrumental planning and trial initiation; Ali Bohra (Orphan Reach, CRO) for spearheading the trial locally in India.

## Author contributions

Conception and design: G. Tornling, R. Batta, J. Porter, T. Bengtsson, A. Hallberg, C. -J. Dalsgaard, J. Raud. Data acquisition: R. Batta, K. Parmar, R. Kashiva, A.K. Cohrt, K. Westergaard. Analysis and interpretation: all authors. Drafting of the manuscript: G. Tornling, R. Batta, C. -J. Dalsgaard, J. Raud. Manuscript review and approval: all authors.

