## Supplement for "The angiotensin type 2 receptor agonist C21 restores respiratory function in COVID19 - a double-blind, randomized, placebo-controlled Phase 2 trial"

### Investigators in the ATTRACT trial

| **Coordinating Investigator** | |
|---|---|
| Professor Joanna Porter | Department of Thoracic Medicine, University College London Hospital NHS Trust, London NW1 2PG, England & UCL Respiratory |
| **Principle Investigators** | **Investigatory Site** |
| Dr Reema Kashiva | Department of Medicine, Noble Hospitals Pvt. Ltd, 153 Magarpatta City Road, Hadapsar, Pune, Maharashtra, India 411013 |
| Dr Chirag Chhatwani | Infectious Disease, Metas Adventist Hospital, 13-B, Ninth No 0363 to 0365, RS No 21, Opp. Chowpati Main Road, Athwalines, Surat, Gujarat, India 395001 |
| Dr Atul Rajkondawar | Department of Medicine, Government Medical College and Hospital, Hanuman Nagar, Ajni Rd, Medical Chowk, Ajni, Nagpur, Maharashtra, India 440003 |
| Dr Paritosh Baghel | Internal Medicine S.L. Raheja Hospital Raheja Rugnalaya Marg, Mahim (W), Mumbai, Maharashtra. India 400016 |
| Dr Kartikeya Parmar | Medicine Department, Civil Hospital and B J Medical College, Asarwa, Ahmedabad, Gujarat, India 380016 |
| Dr Kapil Zirpe | "Neuro Critical Care, Grant Medical Foundation Ruby Hall Clinic, 40 Sasoon Rd, Sangamvadi, Pune, Maharashtra, India 411001 |
| Dr. Mahesh Sutariya | Critical Care Medicine, Unity Trauma Center and ICU (Unity Hospital) Nr. D.R. World, Opp Raghuvir Business Empire, Aai Mata Rd, Parvat Patiya, Surat, Gujarat, India 395010 |
| Dr Nirav Bhalani | Clinical Research Department, Near Siddharth Bunglows, Sama-Savli Road, Vadodara, Gujarat, lndia 390022 |

### Inclusion and Exclusion Criteria in the ATTRACT trial

**Inclusion Criteria**

1) Written informed consent, consistent with ICH GCP R2 and local laws, obtained before the initiation of any trial related procedure
2) Diagnosis of coronavirus (SARS-CoV)-2 infection confirmed by polymerase chain reaction (PCR) test < 4 days before Visit 1 with signs of an acute respiratory infection
3) Age > 18 and < 70 years
4) CRP > 50 and < 150 mg/l
5) Admitted to a hospital or controlled facility (home quarantine is not sufficient)
6) In the opinion of the Investigator, the subject will be able to comply with the requirements of the protocol

**Exclusion Criteria**

1) Any previous experimental treatment for COVID-19
2) Need for mechanical invasive or non-invasive ventilation
3) Concurrent respiratory disease such as chronic obstructive pulmonary disease, idiopathic pulmonary disease (IPF) and/or intermittent, persistent or more severe asthma requiring daily therapy or any subjects that have had an asthma flare requiring corticosteroids in the 4 weeks (28 days) prior to COVID-19 diagnosis
4) Participation in any other interventional trial within 3 months prior to Visit 1
5) Any of the following findings at Visit 1:
   - Positive results for hepatitis B surface antigen (HBsAg), hepatitis C virus antibody (HCVAb) or human immunodeficiency virus 1+2 antigen/antibody (HIV 1+2 Ag/Ab)
   - Positive pregnancy test (see Section 8.2.3)
6) Clinically significant abnormal laboratory value at Visit 1 indicating a potential risk for the subject if enrolled in the trial as evaluated by the investigator
7) Concurrent serious medical condition with special attention to cardiac or ophthalmic conditions (e.g. contraindications to cataract surgery), which in the opinion of the Investigator makes the subject inappropriate for this trial
8) Malignancy within the past 3 years with the exception of in situ removal of basal cell carcinoma and cervical intraepithelial neoplasia grade I
9) Treatment with any of the medications listed below within 1 week prior to Visit 1:
   - Strong Cytochrome p450 (CYP) 3A4 inducers (e.g. rifampicin, phenytoin, St. John’s Wort, phenobarbital, rifabutin, carbamazepine, anti HIV drugs, barbiturates)
   - Warfarin
10) Pregnant or breast-feeding female subjects
11) Female subjects of childbearing potential not willing to use contraceptive methods as described in Section 5.3.1
12) Male subjects not willing to use contraceptive methods as described in Section 5.3.1
13) Subjects known or suspected of not being able to comply with this trial protocol (e.g. due to alcoholism, drug dependency or psychological disorder)

### Withdrawal Criteria in the ATTRACT trial

| |
|---|
| *A subject will be withdrawn from IMP if any of the following occurs:*<br>• Need for mechanical invasive or non-invasive ventilation<br>• Discharge from the hospital/controlled facility<br>• Major protocol deviations as defined by Sponsor.<br>• Sponsor decision to stop the trial or to stop the subject's participation in the trial; reasons may include medical, safety, or regulatory issues, or other reasons consistent with applicable laws, regulations, and GCP.<br>• It is the wish of the subject for any reason<br>• The Investigator judges it necessary due to medical reasons<br>• Adverse events such as:<br> - Serious cardiovascular complications such as severe peripheral oedema or significant bradycardia indicating a potential risk for the subject as evaluated by the Investigator<br> - Moderate to severe skin rashes as judged by the Investigator e.g. Stevens-Johnson syndrome and toxic epidermal necrolysis<br>• Pregnancy<br>*Withdrawal from trial:*<br>• Enrolment in other clinical studies involving investigational products or enrolment in other types of clinical research judged not to be scientifically or medically compatible with this trial.<br>• Disallowed treatment during the trial period<br>• It is the wish of the subject for any reason<br>• The Investigator judges it necessary due to medical reasons<br>• Adverse events<br>• Lost to follow-up |

### Endpoints in the ATTRACT trial

| |
|---|
| **Primary endpoint**<br>Change from baseline in C-reactive protein (CRP) after treatment with C21 200 mg daily dose (100 mg b.i.d.) |
| **Secondary endpoints**<br>1) Change from baseline in:<br> - Body temperature<br> - IL-6<br> - IL-10<br> - TNF<br> - CA125<br> - Ferritin<br>2) Number of subjects not in need of oxygen supply<br>3) Number of subjects not in need of mechanical invasive or non-invasive ventilation<br>4) Time to need of mechanical invasive or non-invasive ventilation<br>5) Time on oxygen supply (for those not needing mechanical invasive or non-invasive ventilation<br>6) Adverse events |
| **Exploratory endpoints**<br>Blood samples will be saved for potential future analyses of biomarkers reflecting inflammation and lung injury |

### Table E1. Effect on biomarkers

| Biomarker | C21 (N=51) | Placebo (N=55) | Treatment Ratio |
|---|---|---|---|
| CRP - all patients (mg/L) | | | |
| Screening | 86.0 (30.2) | 80.1 (26.5) | |
| Baseline | 49.8 (38.7) | 61.5 (47.6) | |
| End of treatment | 13.2 (14.7) | 23.9 (35.6) | |
| Adjusted treatment geometric mean baseline ratio | 0.19 | 0.22 | 0.85 (p=0.489) |
| CRP - patients with $O_2$ treatment at baseline (mg/L) | | | |
| Baseline | 41.4 (31.9) | 65.1 (51.4) | |
| End of treatment | 12.5 (13.3) | 25.4 (37.3) | |
| Adjusted treatment geometric mean baseline ratio | 0.16 | 0.28 | 0.59 (p=0.088) |
| Interleukin 10 (pg/mL) | | | |
| Baseline | 8.41 (8.90) | 10.06 (12.81) | |
| End of treatment | 13.12 (35.51) | 8.08 (12.48) | |
| Adjusted treatment geometric mean baseline ratio | 0.66 | 0.73 | 0.90 (p=0.535) |
| Interleukin 6 (pg/mL) | | | |
| Baseline | 51.13 (114.41) | 34.85 (39.79) | |
| End of treatment | 59.39 (155.97) | 27.83 (60.95) | |
| Adjusted treatment geometric mean baseline ratio | 0.73 | 0.73 | 1.00 (p=0.992) |
| Tumor Necrosis Factor (pg/mL) | | | |
| Baseline | 17.02 (18.70) | 18.29 (16.65) | |
| End of treatment | 24.53 (56.21) | 23.86 (46.59) | |
| Adjusted treatment geometric mean baseline ratio | 0.91 | 1.01 | 0.90 (p=0.474) |
| CA125 (μ/mL) | | | |
| Baseline | 14.28 (12.88) | 20.46 (38.78) | |
| End of treatment | 16.58 (10.86) | 19.72 (18.49) | |
| Adjusted treatment geometric mean baseline ratio | 1.16 | 1.17 | 0.99 (p=0.942) |
| Ferritin (ng/mL) | | | |
| Baseline | 464.2 (324.6) | 707.5 (564.3) | |
| End of treatment | 411.6 (331.3) | 549.4 (550.6) | |
| Adjusted treatment geometric mean baseline ratio | 0.75 | 0.74 | 1.00 (p=0.973) |

Values are Mean (SD) unless otherwise specified.

### Table E2. Need for supplemental oxygen therapy

| Biomarker | C21 (N=51) | Placebo (N=55) | p-value |
|---|---|---|---|
| All subjects (C21 N=51, Placebo N=55) | | | |
| Oxygen therapy at end of treatment, N (%) | 14 (27.5) | 25 (45.5) | 0.055 |
| Oxygen therapy at Day 14, N (%) | 1 (2.0) | 11 (20.0) | 0.003 |
| Days on oxygen up to Day 14, N (%) | 259 (36.3) | 353 (45.8) | 0.190 |
| Subjects on oxygen at baseline (C21 N=37, Placebo N=39) | | | |
| Oxygen therapy at end of treatment, N (%) | 14 (37.8) | 23 (59.0) | 0.065 |
| Oxygen therapy at Day 14, N (%) | 1 (2.7) | 10 (25.6) | 0.004 |
| Days on oxygen up to Day 14, N (%) | 247 (47.7) | 323 (59.2) | 0.101 |
| Subjects not on oxygen at baseline (C21 N=14, Placebo N=16) | | | |
| Oxygen therapy at end of treatment, N (%) | 0 (0.0) | 2 (12.5) | 0.171 |
| Oxygen therapy at Day 14, N (%) | 0 (0.0) | 1 (1.8) | 0.341 |
| Days on oxygen up to Day 14, N (%) | 12 (6.1) | 30 (13.4) | 0.695 |

### Table E3. Treatment-Emergent Adverse Events by MedDRA System Organ Class

| System organ class<br>Preferred term | C21 (N=51) | Placebo (N=55) |
|---|---|---|
| Metabolism and nutrition disorders | 19 (23.5) | 12 (18.2) |
| Hyperglycemia | 14 (21.6) | 5 (7.3) |
| Hyponatremia | 2 (3.9) | 3 (5.5) |
| Dyslipidemia | 1 (2.0) | 2 (3.6) |
| Gastrointestinal disorders | 4 (7.8) | 6 (10.9) |
| Constipation | 0 (0.0) | 4 (7.3) |
| Renal and urinary disorders | 1 (2.0) | 4 (7.3) |
| Glycosuria | 1 (2.0) | 3 (5.5) |

Table includes Adverse Events reported by >2 subject
Data presented as number of events (% of subjects)
