## Supplementary material for "The angiotensin type 2 receptor agonist C21 restores respiratory function in COVID19 - a double-blind, randomized, placebo-controlled Phase 2 trial": Clinical Trial Protocol

| | |
|---|---|
| **Title:** | **A randomised, double-blind, placebo-controlled, phase 2 trial investigating the safety and efficacy of C21 in hospitalised subjects with COVID-19 infection not requiring mechanical ventilation** |
| **Short Title:** | **C21 in COVID-19** |
| **Sponsor:** | **Vicore Pharma AB**<br>**Kronhusgatan 11**<br>**SE-411 05 Göteborg**<br>**Sweden** |
| **Trial ID:** | **VP-C21-006** |
| **EudraCT No.:** | **2020-001502-38** |
| **Investigational Medicinal Product:** | **C21 (Compound 21)** |
| **Indication:** | **Coronavirus disease (COVID-19; SARS CoV-2) infection** |
| **Phase:** | **2** |
| **Version:** | **2.0** |
| **Date:** | **14-Aug-2020** |

Angiotensin II Type Two Receptor Agonist COVID-19 Trial

#### The ATTRACT Trial

*This Clinical Trial Protocol is the property of Vicore Pharma AB and is a confidential document. It is not to be copied or distributed to other parties without written approval from Vicore Pharma AB.*

#### Sponsor's Approval of Clinical Trial Protocol

This trial protocol was subject to critical review by the Sponsor. The information it contains is consistent with current knowledge of the risks and benefits of the investigational medicinal product, as well as with the ethical and scientific principles governing clinical research as set out in the latest version of the Declaration of Helsinki and the International Conference on Harmonisation (ICH) Guideline for Good Clinical Practice (ICH-GCP R2).

This trial will be conducted in compliance with the protocol, GCP, and applicable regulatory requirements.

| | | |
|---|---|---|
| **Sponsor's Medical Responsible:** | Rohit Batta (Aug 14, 2020 14:18 GMT+1) | 14-Aug-2020 |
| Chief Medical Officer | | |
| Rohit Batta, MBBS MRCGP FFPM | Signature | Date |
| Vicore Pharma AB | | |
| **Sponsor's Statistical Expert:** | Thomas Bengtsson<br>Thomas Bengtsson (Aug 14, 2020 16:31 GMT+2) | 14-Aug-2020 |
| Thomas Bengtsson, M.Sc | Signature | Date |
| StatMind AB | | |
| **VP Clinical Development:** | Mimi Flensburg | 14-Aug-2020 |
| Mimi F. Flensburg, DVM, PhD | Signature | Date |
| Vicore Pharma AB | | |

#### Signatory Investigator's Approval of Clinical Trial Protocol

This trial will be conducted in compliance with the protocol, GCP, and the applicable regulatory requirements.

I confirm, that I agree to conduct this trial in compliance with the Declaration of Helsinki, the International Conference on Harmonisation (ICH) Guideline for GCP (ICH-GCP R2) and applicable regulatory requirements.

Furthermore, I confirm that I have read and understood the present clinical trial protocol and agree to conduct the trial in compliance with this. I fully understand that any changes from the clinical trial protocol constitute a deviation which will be notified to Sponsor.

**Coordinating Investigator:**

Joanna C Porter (Aug 23, 2020 21:59 GMT+1)

Signature

23-Aug-2020

Date

Professor Joanna Porter,

MA MB BCh FRCP PhD

Department of Thoracic Medicine

250 Euston Road

London NW1 2PG

19.3 Publication of Results 43
20 INSURANCE AND LIABILITY 43
21 REFERENCES 44
22 ABBREVIATIONS 48

#### 1 PROTOCOL SUMMARY

| | |
|---|---|
| **Trial Title** | A randomised, double-blind, placebo-controlled, phase 2 trial investigating the safety and efficacy of C21 in hospitalised subjects with COVID-19 infection not requiring mechanical ventilation |
| **Trial ID** | VP-C21-006 |
| **Trial p** | 2 |
| **Objectives** | **Primary objective**<br>To investigate the efficacy of C21 200 mg daily dose (100 mg *b.i.d.*) on COVID-19 infection not requiring mechanical invasive or non-invasive ventilation<br><br>**Secondary objectives**<br>To evaluate the following of C21 200 mg daily dose (100 mg *b.i.d.*)<br>• Effect on inflammation<br>• Safety profile<br><br>**Exploratory objectives**<br>To investigate a range of laboratory parameters as potential biomarkers of inflammation and viral load, following oral administration of C21 200 mg daily dose (100 mg *b.i.d.*). |
| **Endpoints** | **Primary endpoint**<br>• Change from baseline in C-reactive protein (CRP) after treatment with C21 200 mg daily dose (100 mg *b.i.d.*)<br><br>**Secondary endpoints**<br>1) Change from baseline in:<br>• Body temperature<br>• IL-6<br>• IL-10<br>• TNF<br>• CA125<br>• Ferritin<br>2) Number of subjects not in need of oxygen supply<br>3) Number of subjects not in need of mechanical invasive or non-invasive ventilation<br>4) Time to need of mechanical invasive or non-invasive ventilation<br>5) Time on oxygen supply (for those not needing mechanical invasive or non-invasive ventilation<br>6) Adverse events<br><br>**Exploratory endpoints**<br>Blood samples will be saved for potential future analyses of biomarkers reflecting inflammation and lung injury |

| | |
|---|---|
| **Trial Design** | A randomised, double-blind, placebo-controlled trial evaluating the efficacy and safety of C21 200 mg daily dose (100 mg *b.i.d)* on top of standard of care for 7 days.<br><br>Approximately 150 subjects with COVID-19 infection will be randomised 1:1 to receive either standard of care + C21 (N=75) or standard of care + placebo (N=75).<br><br>All subjects will be followed-up 7-10 days after receiving the last investigational medicinal product (IMP) dose (visit or phone call, if recovering at home) |
| **Inclusion Criteria** | 1) Written informed consent, consistent with ICH GCP R2 and local laws, obtained before the initiation of any trial related procedure<br>2) Diagnosis of coronavirus (SARS-CoV)-2 infection confirmed by polymerase chain reaction (PCR) test $\leq$ 4 days before Visit 1 with signs of an acute respiratory infection<br>3) Age $\geq$ 18 and $\leq$ 70 years<br>4) CRP $\geq$ 50 and $\leq$ 150 mg/l<br>5) Admitted to a hospital or controlled facility (home quarantine is not sufficient)<br>6) In the opinion of the Investigator, the subject will be able to comply with the requirements of the protocol |
| **Exclusion Criteria** | 1) Any previous experimental treatment for COVID-19<br>2) Need for mechanical invasive or non-invasive ventilation<br>3) Concurrent respiratory disease such as chronic obstructive pulmonary disease, idiopathic pulmonary disease (IPF) and/or intermittent, persistent or more severe asthma requiring daily therapy or any subjects that have had an asthma flare requiring corticosteroids in the 4 weeks (28 days) prior to COVID-19 diagnosis<br>4) Participation in any other interventional trial within 3 months prior to Visit 1<br>5) Any of the following findings at Visit 1:<br>o Positive results for hepatitis B surface antigen (HBsAg), hepatitis C virus antibody (HCVAb) or human immunodeficiency virus 1+2 antigen/antibody (HIV 1+2 Ag/Ab)<br>o Positive pregnancy test (see Section 8.2.3)<br>6) Clinically significant abnormal laboratory value at Visit 1 indicating a potential risk for the subject if enrolled in the trial as evaluated by the investigator<br>7) Concurrent serious medical condition with special attention to cardiac or ophthalmic conditions (e.g. contraindications to cataract surgery), which in the opinion of the Investigator makes the subject inappropriate for this trial<br>8) Malignancy within the past 3 years with the exception of *in situ* removal of basal cell carcinoma and cervical intraepithelial neoplasia grade I<br>9) Treatment with any of the medications listed below within 1 week prior to Visit 1:<br>o Strong Cytochrome p450 (CYP) 3A4 inducers (e.g. rifampicin, phenytoin, St. John's Wort, phenobarbital, rifabutin, carbamazepine, anti HIV drugs, barbituates)<br>o Warfarin<br>10) Pregnant or breast-feeding female subjects<br>11) Female subjects of childbearing potential not willing to use contraceptive methods as described in Section 5.3.1<br>12) Male subjects not willing to use contraceptive methods as described in Section 5.3.1<br>13) Subjects known or suspected of not being able to comply with this trial protocol (e.g. due to alcoholism, drug dependency or psychological disorder) |

| | |
|---|---|
| **Withdrawal Criteria** | A subject will be withdrawn from IMP if any of the following occurs:<br>• Need for mechanical invasive or non-invasive ventilation<br>• Discharge from the hospital/controlled facility<br>• Major protocol deviations as defined by Sponsor.<br>• Sponsor decision to stop the trial or to stop the subject's participation in the trial; reasons may include medical, safety, or regulatory issues, or other reasons consistent with applicable laws, regulations, and GCP.<br>• It is the wish of the subject for any reason<br>• The Investigator judges it necessary due to medical reasons<br>• Adverse events such as:<br> ○ Serious cardiovascular complications such as severe peripheral oedema or significant bradycardia indicating a potential risk for the subject as evaluated by the Investigator<br> ○ Moderate to severe skin rashes as judged by the Investigator e.g. Stevens-Johnson syndrome and toxic epidermal necrolysis<br>• Pregnancy<br><br>Withdrawal from trial:<br>• Enrolment in other clinical studies involving investigational products or enrolment in other types of clinical research judged not to be scientifically or medically compatible with this trial.<br>• Disallowed treatment during the trial period<br>• It is the wish of the subject for any reason<br>• The Investigator judges it necessary due to medical reasons<br>• Adverse events<br>• Lost to follow-up |
| **IMP** | IMP will be delivered as 50 mg capsules packed in plastic container units with 28 capsules in each.<br><br>Each unit will contain either C21 or matching placebo.<br><br>At the trial site, IMP must be stored separately from normal clinic stocks in a securely locked area only accessible to authorised trial personnel.<br><br>Labeling of the IMP will be in local language and done in compliance with GMP Annex 13 (GMP 2003) and local regulatory requirements.<br><br>IMP will be administered twice daily to the subjects for 7 days as follows:<br>• Morning dose: Two 50 mg capsules (100 mg C21 or placebo) to be taken with a glass of water after minimum 2 hours fasting<br>• Afternoon/evening dose: Two 50 mg capsules (100 mg C21 or placebo) to be taken with a glass of water after minimum 2 hours fasting<br><br>Subjects will be required not to eat anything for 1 hour after taking the IMP. |
| **Allowed Medications** | Concomitant medication may be given according to local standard of care. |
| **Disallowed Medications** | 1 week before Visit 1 and during the trial period<br>• Strong CYP3A4 inducers (e.g. rifampicin, phenytoin, St. John's Wort, phenobarbital, rifabutin, carbamazepine, anti HIV drugs, barbituates)<br>• Warfarin<br>• Experimental drugs |

### 2 FLOW CHARTS

**Table 1** Flow Chart for Trial Procedures

| | **Screening** | **Treatment Period** | | | | | | | **With-drawal[1)]** | **End-of-Trial [2)]** |
|---|---|---|---|---|---|---|---|---|---|---|
| **Visit Number** | **Visit 1** | **Visit 2** | **Visit 3** | **Visit 4** | **Visit 5** | **Visit 6** | **Visit 7** | **Visit 8** | - | **Visit 9** |
| Visit window (days allowed from previous visit) | | 1-3 | 1 | 1 | 1 | 1 | 1 | 1 | - | 7-10[2)] |
| **Eligibility / General** | | | | | | | | | | |
| Informed consent | x | | | | | | | | | |
| Check eligibility criteria | x | x[3)] | | | | | | | | |
| Demographics | x | | | | | | | | | |
| Medical history | x | | | | | | | | | |
| Randomisation | | x | | | | | | | | |
| Supplemental $O_2$ | x | x | x | x | x | x | x | x | | x |
| Previous/concomitant medication | x | x | x | x | x | x | x | x | x | x |
| Check of withdrawal criteria | | x | x | x | x | x | x | x | | |
| **Clinical Safety** | | | | | | | | | | |
| Physical examination[4)] | x[4)] | x | x | x | x | x | x | x | | |
| Body weight | x | | | | | | | | | |
| Height | x | | | | | | | | | |
| Body temperature | x | x | x | x | x | x | x | x | | |
| Vital signs | x | x | x | x | x | x | x | x | | |
| 12-lead ECG[5)] | x | | | | | | | | | |
| Adverse events | x | x | x | x | x | x | x | x | x | x |
| **IMP** | | | | | | | | | | |
| IMP administration[6)] | | x | x | x | x | x | x | x | | |
| Check of fasting criteria | | x | x | x | x | x | x | x | | |
| Drug accountability | | | | | | | | x | x | |

1) Additional withdrawal asessments must be performed at Visit 2-8 if the subject is prematurely withdrawn from IMP (see Section 6.5.3)
2) An end-of-trial visit for all randomised subjects who received IMP must be performed 7-10 days after the last dose of IMP. The end-of-trial visit must also be performed if a subject is withdrawn from the trial for any reason.
3) Re-evaluation of in-/exclusion criteria including evaluation of ECG and laboratory results from Visit 1
4) A complete physical examination will be performed at Visit 1. At Visits 2-8, only a short physical examination will be performed (see Section 8.2.4)
5) All ECGs will be reviewed locally
6) IMP is administered twice daily (see Section 7.4)

**Table 2** Flow Chart for Laboratory Assessments

| **Visit** | **Screening** | **Treatment Period** | | | | | | | **With-drawal**[1] | **End-of-Trial** [2] |
|---|---|---|---|---|---|---|---|---|---|---|
| **Visit Number** | **Visit 1** | **Visit 2** | **Visit 3** | **Visit 4** | **Visit 5** | **Visit 6** | **Visit 7** | **Visit 8** | **-** | **Visit 9** |
| Visit window (days allowed from previous visit) | | 1-3 | 1 | 1 | 1 | 1 | 1 | 1 | - | 7-10 |
| **Laboratory Sampling** | | | | | | | | | | |
| HCV Ab | x | | | | | | | | | |
| HBsAg | x | | | | | | | | | |
| HIV 1+2 Ag/Ab | x | | | | | | | | | |
| Biochemistry[7] | x | x | x | x | x | x | x | x | | |
| Haematology[8] | x | x | x | x | x | x | x | x | | |
| CRP | x | x | x | x | x | x | x | x | x | |
| IL-6 | | x | x | x | x | x | x | x | | |
| IL-10 | | x | x | x | x | x | x | x | | |
| TNF | | x | x | x | x | x | x | x | | |
| CA125 | | x | x | x | x | x | x | x | | |
| Ferritin | | x | x | x | x | x | x | x | | |
| Urinalysis [9] | x | x | x | x | x | x | x | x | | |
| Biomarkers | | x | | | | | | x | x | |
| Pregnancy testing [10] [11] | x | | | | | | | x | x | |

7) Haematology: haemoglobin, haematocrit (erythrocyte volume fraction), platelet count (thrombocyte particle concentration [TPC]), leucocyte count, mean corpuscular volume (MCV), white blood cells with differential count (see Section 8.2.7)
8) Biochemistry: albumin, alanine transferase (ALT), aspartate aminotransferase (AST), alkaline phosphatase (ALP), bilirubin, blood urea nitrogen (BUN), calcium, creatinine, glucose, potassium, sodium (see Section 8.2.7)
9) Urinalysis: dipstick for bilirubin, glucose, ketones, protein, specific gravity, urobilinogen, and pH (see Section 8.2.7)
10) Only applicable for female subjects of childbearing potential, as defined in Section 5.3.1.
11) Urine dipstick is performed – if positive for pregnancy this is to be followed up by a pregnancy blood test

### 3 BACKGROUND AND RATIONALE

#### 3.1 Indication Coronavirus Disease (COVID-19)

Coronavirus disease (COVID-19) is a viral infection caused by the newly emergent coronavirus SARS-CoV-2 (World Health Organization [WHO] In January 2020, SARS-CoV-2 was identified as the causative agent of an outbreak of viral pneumonia centered around Wuhan, Hubei, China, where the first cases were reported in December 2019 (Wu et al., 2020; Zhou et al., 2020; Lu et al., 2020). The virus caused a widespread outbreak of infection throughout China. It rapidly expanded globally and by March 12, 2020, COVID-19 was announced a pandemic by the World Health Organization (WHO).

SARS-CoV-2 is currently thought to originate from a bat host (Zhou et al., 2020; Lu et al., 2020), probably with an intermediate animal host passing the virus from bats to humans (Lake et al., 2020). Human-to-human transmission is now well established and the transmission occurs via the respiratory route or via contact with infected secretions. The basic reproduction number, $R_0$ (the expected number of secondary cases produced by a single [typical] infection in a completely susceptible population), is currently estimated by the WHO as 1.4-2.5 (Phan et al., 2020). Based on 425 cases identified in early January 2020 in Wuhan, the mean incubation period was estimated to be 5.2 days (Li et al., 2020).

COVID-19 infection can cause mild to severe respiratory illness, and symptoms can include fever, cough, and shortness of breath (WHO 2020). As of April 1., 2020, the WHO has reported 823,626 confirmed cases of COVID-19 and 40,598 deaths globally. At present, the European region is the region that is most severely affected with 464,212 confirmed cases of COVID-19 and 30,089 deaths.

Most reported cases of COVID-19 illness are mild, but severe disease is not uncommon. Currently, data are limited but early studies from China have shown that severity was related to the burden of comorbidities including diabetes, hypertension, and cardiovascular disease as well as to increasing age (WHO 2020; Huang et al., 2020). Dysregulation of the renin-angiotensin system (RAS) cascade is present in each of these comorbidities and could increase the likelihood or severity of SARS-CoV-2 infection (Hanff et al., 2020).

Estimates of case fatality ratio (CFR) of COVID-19 are uncertain. In an early trial including 1,975 confirmed cases in China, the CFR was 2.8% (Wang et al, 2020). In a new trial including 70,117 cases in China, a CFR of 3.67% was estimated (Verity et al., 2020). In the same trial, the authors made a best estimate of the CFR in China of 1.38% after adjusting for demography and under-ascertainment and found substantially higher CFRs in age groups above 60 years than below 60 years.

Initial symptoms are not necessarily predictive of disease severity. Several very recent publications have shown that CRP is an indicator of COVID-19 progression that predicts severe disease (e.g. Li et al., 2020; Tan et al., 2020; Wang et al, 2020) and mortality (Ruan et al., 2020).

##### 3.2 Current Treatment for COVID-19

Currently, there is no cure or preventive vaccine for COVID-19. Several clinical trials are ongoing and investigating e.g. antiviral and immunomodulatory agents in the search for urgent, safe, and effective treatment of COVID-19 infection as well as vaccines for prophylactic treatment of COVID-9 infection (WHO). April 1., 2020, 260 trials investigating COVID-19 are listed at www.clinicaltrials.gov. The standard of care is primarily organ support depending on disease severity and complications (ICNARC 2020).

##### 3.3 Investigational Medicinal Product

The investigational medicinal product (IMP) Compound 21 (C21), is a potent and selective angiotensin II (AngII) type 2 receptor ($AT_2R$) agonist presented as 50 mg capsules for oral administration.

C21 is a first-in-class, low molecular weight, orally available high affinity $AT_2R$ agonist in development for COVID-19 and interstitial lung diseases, including idiopathic pulmonary fibrosis (IPF). The $AT_2R$, which can be viewed as an activator of a tissue-protective natural arm of the human body, is mainly expressed in embryonic tissue and, under normal conditions, only at low levels in most tissues in healthy adults (Steckelings et al., 2005; de Gasparo et al., 2002). However, the $AT_2R$ is relatively highly expressed in the adult human lung (Human Protein Atlas), and can be upregulated during repair and regeneration (see e.g. Akishita et al., 1999).

The Renin Angiotensin System (RAS) is an important hormone system that regulates blood pressure, fluid, and electrolyte balance, and several other physiological processes. In the RAS cascade, the hormone angiotensinogen is converted to angiotensin I (AngI) by the enzyme renin, and AngI is then converted to the powerful vasoconstrictor AngII by angiotensin-converting enzyme (ACE). AngII functions through 2 specific receptor types, Angiotensin II Type 1 Receptor ($AT_1R$) and $AT_2R$ which have opposing roles (Azushima et al., 2020).

The $AT_1R$ mediates the classical actions of AngII, including constriction of blood vessels, sodium retention, and cell growth. However, abnormal $AT_1R$ activation contributes to the pathogenesis of cardiovascular and renal diseases such as hypertension, myocardial infarction, heart failure, different fibrotic diseases as well as the “cytokine storm” causing acute respiratory distress syndrome (ARDS) in COVID-19 (Henderson et al., 2020). Conversely, activation of $AT_2R$ causes dilatation of blood vessels and inhibition of inflammation, apoptosis, and fibrosis, and is thus considered to play a beneficial counter-regulatory role to the effects of $AT_1R$ activation (Matavelli and Siragy, 2015). The natural ligands/agonists of $AT_2R$, Ang 1-7 and Ang 1-9, are the products of angiotensin-converting enzyme 2 (ACE2), a homologue to ACE, cleaving AngII and AngI, respectively (Guignabert et al., 2018). Interstingly, both ACE2 and $AT_2R$ have been shown to protect mice from severe acute lung injury induced by acid aspiration or sepsis (Imai et al., 2005), and C21 has been shown to significantly reduce pulmonary inflammation in different animal models of acute lung injury by reducing pro-inflammatory cytokines such as IL-6 and TNF (Bruce et al., 2015; Menk et al., 2018; Rathinasabapathy et al., 2018).

ACE2 has been identified as the functional SARS-CoV receptor for cellular entry and replication, both *in vitro* (Li et al., 2003) and *in vivo* (Kuba et al., 2005). It is also known that such infection results in downregulation of ACE2 (Zhang et al., 2020). Because ACE2 protects against inflammatory lung injury, the SARS-CoV-mediated ACE2 downregulation is believed to contribute to virus-induced lung pathologies (Kuba et al., 2005) by a reduction of inhibitory control over toxic AngII and several pathological complications such as ARDS. More recently, it was shown that ACE2 is also the cellular receptor for SARS-CoV-2, the COVID-19 pathogen (Zhou et al., 2020), and it has been suggested that the interaction between ACE2 and CoV-1/CoV-2 (which is not seen with other less harmful corona viruses) may explain the shift in corona virus morbidity and mortality.

Taken together, Vicore's findings and the fact that the RAS plays a key role in the development of COVID-19 suggest that C21 may have a role in the treatment of the disease by directly activating $AT_2R$ downstream of ACE2 and thus counteracting the deranged ACE2/ACE imbalance in the infected state.

##### 3.4 Preclinical Safety

C21 has undergone an extensive nonclinical safety and toxicology evaluation including safety pharmacology, single, and repeated dose toxicity up to 13 weeks in rat, dog, and, cynomolgus monkey, genotoxicity, and phototoxicity studies. The key findings from the safety pharmacology studies and toxicology studies are presented in Table 3.

**Table 3 Summary of Key Safety and Toxicology Findings**

| | **Rat** | | **Dog** | | **Cynomolgus Monkey** | |
|---|---|---|---|---|---|---|
| **Findings** | **Effect Dose (mg/kg)** | **No Effect Dose (mg/kg)** | **Effect Dose (mg/kg)** | **No Effect Dose (mg/kg)** | **Effect Dose (mg/kg)** | **No Effect Dose (mg/kg)** |
| Increased blood pressure | n.a. | n.a. | 1[1)] | n.o. | n.o | 30 |
| Increased QTcR interval | n.a | n.a | 25[1)] | 15 | n.o | 30 |
| Increased PR interval | n.a | n.a | 10[1)] | 1 | n.o | 30 |
| Lens opacities | 20 | 6 | n.o. | 25 | n.o | 30 |

n.a., not analysed; n.o., not observed; [1)]only observed in single dose telemetry trial.

In the rat, the main toxicological finding was an irreversible uni- or bilateral eye opacity after 13 weeks of treatment with C21 at doses of 20 and 60 mg/kg/day. The lenticular changes seemed to progress after cessation of the treatment. Lens opacity or lenticular changes were not observed in either dog or cynomolgus monkey and are therefore thought to be specific to the rat species.

Other findings observed in rats after treatment of C21 at doses up to 60 mg/kg/day for 4-13 weeks included reversible histopathological changes and/or higher organ weights of the liver, the adrenal, the ovary, the pituitary, and the thyroid gland. The no observed adverse effect level (NOAEL) in rats was determined to be 6 mg/kg/day in the 13-week study.

In the dog, daily doses of 30 mg/kg/day C21 for 14 days were tolerated but were associated with small reductions in food intake and body weight, salivation, and emesis. In a 4-week toxicity study, no signs of toxicity were detected at doses up to 25 mg/kg/day but increases in blood pressure were observed in males administered 25 mg/kg/day. In the 13-week study, only marginal increases in blood pressure were observed after administration of 15 mg/kg/day and these were considered non-adverse as all values remained within the normal biological variation range.

Other findings in dogs after administration of 15-25 mg/kg/day for up to 13 weeks included small reversible changes in faecal consistency, decreased food consumption and an associated reduction in body weight gain, mild haemoconcentration (both sexes), slightly elevated monocyte count (males) and fibrinogen levels (both sexes), increased overnight urine volumes and an associated reduction in urine specific gravity (females).

In safety pharmacology studies, cardiovascular effects of C21 treatment were observed in Beagle dogs. The main findings were increased blood pressure and electrocardiography (ECG) aberrations. Increases in systolic and diastolic arterial pressure were observed at doses of 1 to 25 mg/kg in a single dose telemetry study and in a 4-week toxicity study only at the highest dose of 25 mg/kg/day. Effects on ECG were observed at single doses of 10 and 25 mg/kg in the dog telemetry study and consisted of PR and QT/QTcR (QT interval with Rautaharju's correction) prolongations, atrioventricular block I and II, and isolated premature ventricular contractions. These ECG aberrations could well be explained by the increase in blood pressure. In a 4-week dog toxicity study of C21 (up to 25 mg/kg/day), no ECG changes were observed.

In cynomolgus monkeys, treatment with 100 mg/kg/day C21 for 5 days resulted in emesis, slightly reduced body weight, excessive salivation, and liquid faeces. No test item related-findings were observed during the in-life ophthalmic investigations or after microscopic evaluation of the eyes at doses up to 30 mg/kg/day after 13 weeks of dosing. Administration of C21 at dose levels of 10-30 mg/kg/day in cynomolgus monkey was associated with excessive salivation, liquid faeces, and isolated areas of squamous skin. In addition, an increase in absolute and relative liver weight was observed in males, however this effect was probably a secondary effect due to hepatic P450 induction. The NOAEL in cynomolgus monkey was determined to be 30 mg/kg/day in the 13-week study.

There are no indications of mutagenic, genotoxic or phototoxic potential of C21.

For further details, please refer to the current Investigator's Brochure.

#### 3.5 Clinical Safety

Safety and tolerability of C21 have been evaluated in three phase 1 trials of which 2 trials are completed (C21-001-16, C21-002-16) and 1 trial is in the reporting phase (C21-003 [preliminary data]). During these trials, C21 was evaluated in single ascending dose (SAD) and multiple ascending dose (MAD) trials at doses up to 200 mg twice daily for up to 8 days in healthy subjects and at doses up to 100 mg daily for up to 8 days in obese subjects. These trials included 83 subjects receiving at least 1 dose of C21.

C21 was rapidly absorbed in all 3 trials with median $t_{max}$ ranging from 20 to 60 min. The plasma concentration of C21 increased with dose in a dose-dependent (C21-003) or nearly supra-proportional manner (C21-001-16, C21-002-16). Little or no accumulation of C21 was observed after 8 days of dosing. The exposure of C21 was substantially decreased under fed conditions as compared to fasted conditions (C21-003). The maximal plasma concentration ($C_{max}$) and area under the curve ($AUC_{0-\infty}$) were reduced in a statistically significant manner and were 6.5 and 2.3 times lower, respectively, in the fed state when compared to the fasted state. In addition, median $t_{max}$ was delayed by approximately 35 min in presence of food. These findings indicate a strong interaction with food.

Levels of the main metabolite of C21 in plasma, M1, were also quantified during the trials. M1 was rapidly formed across the trials and median $t_{max}$ was reached a little later than for C21, ranging from 75 to 150 min. The plasma concentration of M1 also increased with dose in a nearly supra-proportional manner. Little or no accumulation of M1 was observed after 8 days of dosing.

Across the trials, C21 was generally well tolerated at daily doses up to 200 mg, whereas doses of 200 mg twice daily (400 mg daily) for 8 days resulted in reversible alopecia in 6 out of 6 subjects occurring within 2 weeks after end of dosing and improving within 8 weeks after end of dosing (C21-003). The observed alopecia was considered related to C21-treatment and is possibly due to a secondary pharmacological effect of stimulation of the $AT_2R$.

The most frequently reported treatment-emergent adverse events (TEAE) related to C21 treatment was headache. Other TEAEs related to C21 treatment included dizziness, nausea, and fatigue that were reported by 2 subjects and pruritus, rash, vomiting, and diarrhoea that were reported as single events. One incidence of syncope of moderate intensity was reported as being related to C21 treatment (100 mg C21; C21-002-16) and one incidence of presyncope was observed and assessed as related to C21 treatment (100 mg C21; C21-002-16).

One episode of clinically significant PR prolongation was reported in 1 subject's ECG, recorded 1 hour after intake of 3 mg C21 (C21-001-16). This event was assessed as related to C21 treatment. However, a causal relationship with C21 treatment could not be confirmed or excluded and at this stage, it cannot be excluded that C21 could cause slowing of atrioventricular conduction (= PR prolongation) in susceptible individuals, although it is considered unlikely with a documented first and second degree atrioventricular block in the subject 1 month after the treatment period and given the reassuring telemetry findings within the recent phase 1 trial (C21-003) that C21 treatment did not affect ECG values.

There were no clinically significant abnormal findings in the safety laboratory parameters that were considered to be related to C21 treatment and no clinically significant findings or dose-related trends in blood pressure, heart rate, and ECG except for the above mentioned PR prolongation.

No serious adverse events (SAEs) or deaths observed during the 3 phase 1 trials.

In addition, a phase 2a trial in subjects with Raynaud's phenomenon secondary to SSc (SSc-RP) to show proof of mechanism of C21 on vasodilation is currently ongoing.

For further details, please refer to the current Investigator's Brochure.

##### 3.6 Rationale for Trial

There is no cure or effective specific therapeutics for treating patients with COVID-19. A 51% mortality has been reported in the UK for patients in critical care (ICNARC 2020). The standard of care is primarily focussed on organ support such as respiratory failure. A safe and effective treatment is urgently needed to reduce mortality and improve clinical disease outcome such as from ARDS which is thought to manifest from the exuberant cytokine storm associated with the adaptive stage of the disease.

The current clinical trial is the first investigation of C21 in patients infected with COVID-19. Data from the trial will guide and support the design of further clinical investigations of C21 in this indication.

##### 3.7 Rationale for Trial Design

Given the urgent need for effective medicines in COVID-19, proof of concept for C21 is sought within this randomised, double-blind, placebo-controlled phase 2 trial evaluating the safety and efficacy in subjects with COVID-19 infection who have been hospitalised and are not requiring mechanical invasive or non-invasive ventilation. The aim is to intervene early in order to prevent more serious complication such as ARDS and/or mechanical ventilation.

Approximately 150 subjects will be randomised in a 1:1 schedule to receive either standard of care + placebo or standard of care + C21 200 mg daily (100 mg *b.i.d.*[ bis in die; twice daily]) for 7 days.

Rationale for the primary endpoint is supported by several very recent publications that have shown that CRP is an indicator of COVID-19 progression predicting severe disease (e.g. Li et al., 2020; Tan et al., 2020; Wang et al, 2020) and mortality (Ruan et al., 2020).

Data from the trial will guide the design of further clinical investigations of C21 in this indication.

##### 3.8 Rationale for Dose and Dosing Regimen

C21 is being developed for oral treatment of COVID-19. COVID-19 is a viral infection that spreads easily and may cause severe respiratory illness that is potentially life-threatening and represents a huge challenge to the health care system.

The results of the phase 1 healthy subject trials for C21 (C21-003; preliminary data) indicate an acceptable safety and tolerability profile for the 100 mg *b.i.d.* dose. The risks of C21 in adults are considered primarily related to reversible alopecia seen at the higher dose of 200 mg *b.i.d.* In the recent phase 1 trial, the 100 mg *b.i.d.* dose administered for 8 days did not cause any hair loss. Alopecia is therefore not considered to present any significant safety risk to subjects in this trial.

In the recent phase 1 trial (C21-003), 100 mg *b.i.d* dosing prolonged the duration of the plasma concentration at therapeutic relevant levels. After oral administration, C21 was rapidly absorbed. On day 8, median $t_{max}$ was 0.33 hours after each dose post-dose, and C21 was only measurable in plasma until 4-6 hours after the first and second dose. Lower doses than 100 mg *b.i.d.* may not provide maximal efficacy benefit.

The most recent information on the natural course of hospitalised COVID-19 patients, indicate that these patients generally deteriorate within 48 hours and are either discharged or taken into intensive care within 7 days. Hence, it is currently believed, that 7 days of treatment with C21 is a sufficient treatment duration for a conclusive first trial in this population.

Overall, the safety profile of C21 as available from preclinical data and from the phase 1 trials is interpreted as favourable for further study of the intended indication 'Treatment of COVID-19'.

##### 3.9 Risk-Benefit Assessment

Currently, there is no cure or preventive vaccine for COVID-19 and the disease has a mortality rate of 51% for patients in critical care (ICNARC 2020).

The risks to subjects in the present trial will be reduced by using a C21 dosing regimen (100 mg *b.i.d* for 7 days), which has been evaluated in the recently completed phase 1 trial (C21-003) and found to have an acceptable safety profile.

The risks of treatment with C21 are primarily considered related to alopecia. However, in the phase I trial, alopecia occurred only in subjects who received 200 mg *b.i.d.* for 8 days. Regrowth of the hair observed to date provides reassurance that any such an effect is reversed on cessation of treatment.

There is a hypothetical risk of cataract, given lens opacities seen in one species, the rat; however, the dog and the non-human primate showed no evidence at all of this finding, increasing the likelihood that the effect is rat specific. In the recently conducted phase 1 trial (C21-003), no effect was demonstrated on the lens of subjects exposed to 100 mg *b.i.d* or 200 mg *b.i.d* for 8 days.

In the present trial, any potential risks will be minimised by strict compliance with the eligibility criteria. All subjects participating in the trial will be followed closely by medically qualified staff throughout the trial period (see Section 2 and Section 6.1). Adverse events (AEs) will be carefully documented, and the subjects will receive relevant treatment should they occur.

Furthermore, the protocol defines withdrawal criteria (see Section 5.4) to secure the safety of subjects with severe progression of disease. If the physician judges it necessary for the subject to receive mechanical ventilation or a different treatment during the trial period, the subject will be withdrawn from the trial and receive relevant treatment.

Based on the above, it is considered ethically justifiable to include subjects in the present trial, as the potential benefits of C21 are judged to balance possible disadvantages connected with participation in the trial.

#### 4 OBJECTIVES AND ENDPOINTS

##### 4.1 Primary Objective

To investigate the efficacy of C21 200 mg daily dose (100 mg *b.i.d.*) on COVID-19 infection not requiring mechanical invasive or non-invasive ventilation.

##### 4.2 Secondary Objectives

To evaluate the following of C21 200 mg daily dose (100 mg *b.i.d.*)

- Effect on inflammation
- Safety profile

##### 4.3 Exploratory Objectives

To investigate a range of laboratory parameters as potential biomarkers of inflammation and viral load, following oral administration of C21 200 mg daily dose (100 mg *b.i.d.*).

##### 4.4 Primary Endpoint

Change from baseline in C-reactive protein (CRP) after treatment with C21 200 mg daily dose (100 mg *b.i.d.*).

##### 4.5 Secondary Endpoints

1) Change from baseline in:
    - Body temperature
    - IL-6
    - IL-10
    - TNF
    - CA125
    - Ferritin
2) Number of subjects not in need of oxygen supply
3) Number of subjects not in need of mechanical invasive or non-invasive ventilation
4) Time to need of mechanical invasive or non-invasive ventilation
5) Time on oxygen supply (for those not needing mechanical invasive or non-invasive ventilation)
6) Adverse events

##### 4.6 Exploratory Endpoints

Blood samples will be stored for potential future analyses of biomarkers reflecting inflammation and lung injury.

### 5 TRIAL POPULATION

#### 5.1 Inclusion Criteria

1) Written informed consent, consistent with ICH-GCP R2 and local laws, obtained before the initiation of any trial related procedure
2) Diagnosis of coronavirus (SARS-CoV)-2 infection confirmed by polymerase chain reaction (PCR) test ≤ 4 days before Visit 1 with signs of an acute respiratory infection
3) Age ≥ 18 and ≤ 70 years
4) CRP ≥ 50 and ≤ 150 mg/l
5) Admitted to a hospital or controlled facility (home quarantine is not sufficient)
6) In the opinion of the Investigator, the subject will be able to comply with the requirements of the protocol

#### 5.2 Exclusion Criteria

1) Any previous experimental treatment for COVID-19
2) Need for mechanical invasive or non-invasive ventilation
3) Concurrent respiratory disease such as COPD (chronic obstructive pulmonary disease), IPF and/or intermittent, persistent or more severe asthma requiring daily therapy or any subjects that have had an asthma flare requiring corticosteroids in the 4 weeks (28 days) prior to COVID-19 diagnosis
4) Participation in any other interventional trial within 3 months prior to Visit 1
5) Any of the following findings at Visit 1:
   - Positive results for hepatitis B surface antigen (HBsAg), hepatitis C virus antibody (HCVAb) or human immunodeficiency virus 1+2 antigen/antibody (HIV 1+2 Ag/Ab
   - Positive pregnancy test (see Section 8.2.3)
6) Clinically significant abnormal laboratory value at Visit 1 indicating a potential risk for the subject if enrolled in the trial as evaluated by the Investigator
7) Concurrent serious medical condition with special attention to cardiac or ophthalmic conditions (e.g. contraindications to cataract surgery), which in the opinion of the Investigator makes the subject inappropriate for this trial
8) Malignancy within the past 3 years with the exception of *in situ* removal of basal cell carcinoma and cervical intraepithelial neoplasia grade I
9) Treatment with any of the medications listed below within 1 week prior to Visit 1:
   a. Strong Cytochrome p450 (CYP) 3A4 inducers (e.g. rifampicin, phenytoin, St. John's Wort, phenobarbital, rifabutin, carbamazepine, anti HIV drugs, barbituates)
   b. Warfarin
10) Pregnant or breast-feeding female subjects
11) Female subjects of childbearing potential not willing to use contraceptive methods as described in Section 5.3.1

12) Male subjects not willing to use contraceptive methods as described in Section 5.3.1
13) Subjects known or suspected of not being able to comply with this trial protocol (e.g. due to alcoholism, drug dependency or psychological disorder)

#### 5.3 Restrictions During the Trial

##### 5.3.1 Contraception Requirements

Women of child bearing potential must practice abstinence (if that is their preferred lifestyle) from Visit 1 to Visit 9, or must agree to use a highly effective method of contraception with a failure rate of <1% to prevent pregnancy (combined [oestrogen and progestogen containing] hormonal contraception associated with inhibition of ovulation [oral, intravaginal, transdermal], progestogen-only hormonal contraception associated with inhibition of ovulation [oral, injectable, implantable], intrauterine device or intrauterine hormone-releasing system) from at least 4 weeks prior to first IMP administration to 4 weeks after last IMP administration. Their male partner must agree to use a condom during the same time frame, unless he has had a demonstrated successful vasectomy more than 6 months prior to first IMP administration.

Males should use condom and their female partner of child-bearing potential must use a contraceptive method with a failure rate of <1% to prevent pregnancy (see above) and drug exposure of a partner and refrain from donating sperm from the date of dosing until 3 months after the last IMP administration.

##### 5.3.2 Dietary Restrictions

In relation to IMP administrations, subjects should adhere to restrictions regarding food intake:

- No food intake for 2 hours prior to taking the C21 capsules
- No food intake for 1 hour after taking the C21 capsules

#### 5.4 Withdrawal Criteria

In all cases of withdrawal from IMP or trial, the reason(s) for withdrawal, must be recorded in the electronic case report form (eCRF) if known. Note: all attempts should be made to determine the underlying reason for the withdrawal and, where possible, the primary underlying reason should be recorded.

##### 5.4.1 Withdrawal from Trial IMP

If a subject is prematurely withdrawn from IMP, the Investigator must make every effort to perform the withdrawal assessments described in Section 6.5.3, and further perform end-of-trial Visit (Visit 9) 7-10 days after the last dose of IMP.

A subject will be withdrawn from IMP if any of the following occurs:

- Need for mechanical invasive or non-invasive ventilation
- Discharge from the hospital/controlled facility
- Major protocol deviations as defined by Sponsor.

- Sponsor decision to stop the trial or to stop the subject's participation in the trial; reasons may include medical, safety, or regulatory issues, or other reasons consistent with applicable laws, regulations, and GCP.
- It is the wish of the subject for any reason
- The Investigator judges it necessary due to medical reasons
- Adverse events such as:
    - Serious cardiovascular complications such as severe peripheral oedema or significant bradycardia indicating a potential risk for the subject as evaluated by the Investigator
    - Moderate to severe skin rashes as judged by the Investigator e.g. Stevens-Johnson syndrome and toxic epidermal necrolysis
- Pregnancy

##### 5.4.2 Withdrawal from Trial

- Enrolment in other clinical trials involving investigational products or enrolment in other types of clinical research judged not to be scientifically or medically compatible with this trial.
- Disallowed treatment during the trial period
- It is the wish of the subject for any reason
- The Investigator judges it necessary due to medical reasons
- Adverse events
- Lost to follow-up

### 6 TRIAL DESIGN

#### 6.1 Overall Trial Design

This is a randomised, double-blind, placebo-controlled phase 2 trial investigating the safety and efficacy of C21 in subjects who are hospitalised with COVID-19 infection, but not in need of mechanical invasive or non-invasive ventilation.

In total, approximately 150 subjects will be enrolled and randomised to receive twice daily oral administration of either standard of care (SoC) + placebo (N=75) or SoC + C21 (N=75). Subjects will be treated for 7 days.

All subjects will be followed up 7-10 days after receiving the last IMP dose (visit, or phone call if recovering at home).

A total of 9 visits are defined:

- Visit 1: Screening
- Visits 2–8: Treatment period
- Visit 9: End-of-trial

IMP will be administered twice daily (*b.i.d.*) during the treatment period.

Final safety follow-up assessments at Visit 9 (end-of-trial visit), must be performed for all subjects who have received IMP administration. Visit 9 must be scheduled 7-10 days after the last IMP administration (see Section 7.4).

The maximum duration of the trial for any subject will be approximately 3 weeks, including a screening period of up to 3 days, a treatment period of 7 days and a visit for safety follow-up (end-of-trial) to be conducted 7-10 days after the last dose of IMP (see clinical trial design in Figure 1).

#### 6.2 Number of Subjects

The trial will include approximately 150 subjects. Re-screening of subjects failing to meet the in- & exclusion criteria is not allowed.

#### 6.3 Trial Diagram

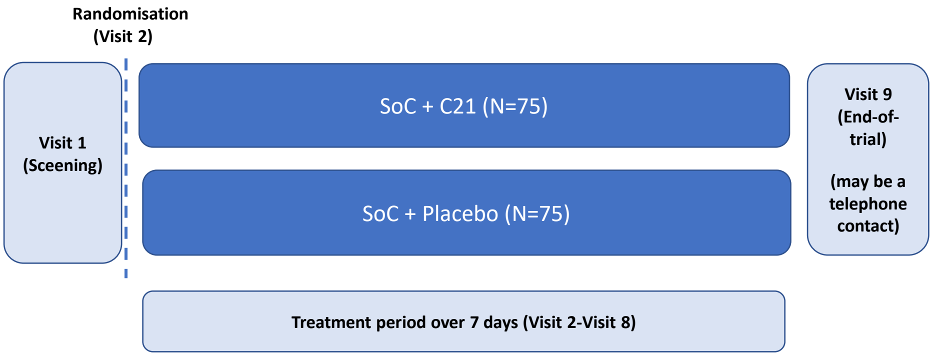

**Figure 1. VP-C21-006 Clinical Trial Design**

#### 6.4 Trial Duration and Participating Centers

Planned first subject screened: Q2 2020

Planned last subject enrolled: Q3 2020

Planned last subject last visit: Q3 2020

The end-of-trial date is defined as the date the last subject completes the last visit.

The trial will be conducted at trial sites  globally.

#### 6.5 Schedule of Events

##### 6.5.1 Screening (Visit 1)

Prior to and no later than at start of screening (Visit 1), i.e. before any trial-related activity takes place, the Investigator or a qualified designee will explain the nature of the trial, its purpose, the procedures involved, the expected duration, the potential risks and benefits involved, and any discomfort the subject may entail. All explanations shall be in layman's language. The subject will be provided with a copy of the information sheet. The subject must be given sufficient time to consider the trial before deciding whether to participate.

After signed informed consent is obtained from the subject, the screening procedures can be initiated.

All subjects giving informed consent to participate in the trial will receive a screening number. During the screening visit, the subject is evaluated for eligibility (see Section 5.1 and 5.2).

The following procedures are performed:

- Written informed consent to be obtained
- Assignment of screening number
- Check of in- & exclusion criteria
- Assessment of eligibility which includes review of demographics, medical history, concomitant illnesses and medication, physical examination, height, body weight, and measurement of blood pressure, pulse, respiration rate, and body temperature (morning values)
- Medical history and concomitant medication
- 12-lead electrocardiography (ECG)
- Recording need of supplementary $O_2$
- Blood sampling for:
    - CRP
    - HBsAG, HCVab, HIV 1+2 Ag/Ab (see Table 2 and Section 8)
    - Safety (haematology and biochemistry) (see Section 8.2.7)
- Urinalysis (dipstick) for bilirubin, glucose, ketones, protein, specific gravity, urobilinogen, pH (see Section 8.2.7)
- Pregnancy testing (urine dipstick), if positive a blood pregnancy test must be performed to confirm positive pregnancy
- Adverse events

##### 6.5.2 Treatment Period (Visits 2-8)

The treatment period lasts for 7 days (Visit 2-8). The procedures to be performed at each visit are specified below.

Visit 2

Visit 2 can be scheduled as soon as the relevant screening assessments are available. Visit 2 must occur within 3 days from Visit 1 (screening).

The following procedures are performed:

- Re-evaluation of in- & exclusion criteria
- Short physical examination (including vital signs)
- Evaluation of ECG and laboratory results from Visit 1
- Assignment of randomisation number
- Concomitant medication
- Recording need of supplementary $O_2$
- Blood sampling for:
  - CRP
  - Safety (haematology and biochemistry) (see Section 8.2.7)
  - IL-6, IL-10, TNF, CA125, ferritin
  - Biomarkers
- Urinalysis (dipstick for bilirubin, glucose, ketones, protein, specific gravity, urobilinogen and pH)
- IMP administration twice daily including diatery restrictions (see Section 5.3.2)
- Check of withdrawal criteria
- Adverse events

Visits 3-8

- Concomitant medication
- Short physical examination (including vital signs at the judgement of the Investigator)
- Recording need of supplementary $O_2$
- Blood sampling for
  - CRP
  - Safety (haematology and biochemistry) (see Section 8.2.7)
  - IL-6, IL-10, TNF, CA125, ferritin
- Urinalysis (dipstick for bilirubin, glucose, ketones, protein, specific gravity, urobilinogen and pH)
- IMP administration twice daily including dietary restrictions (see Section 5.3.2)
- Check of withdrawal criteria
- Adverse events

In addition for Visit 8 (end-of-treatment) only:

- Pregnancy testing (urine dip-stick), if positive a blood pregnancy test must be performed to confirm positive pregnancy
- Biomarkers
- Drug accountability

##### 6.5.3 Withdrawal assessments

If a subject is prematurely withdrawn from IMP at Visit 2-8 (before last dose at Visit 8), additional withdrawal assessments must be performed (see Section 5.4.1). These include:

- Concomitant medication
- Blood sampling for:
    - CRP
    - Biomarkers
- Pregnancy test
- Adverse events
- Drug accountability

##### 6.5.4 End-of-Trial (Visit 9)

Visit 9 is performed for all randomised subjects 7-10 days after the last IMP administration and may be performed as a telephone contact.

The following procedures are performed:

- Concomitant medication
- Date of discharge
- Use of supplemental $O_2$
- Use of mechanical invasive and non-invasive mechanical ventilation
- Adverse events

If a subject is prematurely withdrawn from IMP, Visit 9 must be performed 7-10 days after last IMP dose (see Section 5.4.1).

### 7 TRIAL TREATMENT

#### 7.1 Investigational Medicinal Product

##### 7.1.1 Active Treatment

Active substance: C21 is presented as a capsule for oral administration. The drug substance is 3-[4-(1*H*-imidazol-1-ylmethyl)phenyl]-5-(2-methylpropyl)thiophene-2-[(*N*-butyloxylcarbamate)-sulphonamide] sodium salt supplied as a free form equivalent (eq.) 50 mg C21 oral capsule.

**Table 4 Qualitative and Quantitative Composition of C21**

| Component | Quality reference | Function | Quantity (mg) /unit |
|---|---|---|---|
| C21 (PD00377 Na-salt) | Company specification | Active pharmaceutical ingredient | 52.3[a] |
| Mannitol | Ph. Eur. | Filler | 203.38 |
| Anhydrous colloidal silica | Ph. Eur. | Glidant | 2.14 |
| Magnesium stearate | Ph. Eur. | Lubricant | 2.61 |
| Vcaps® Plus capsule | Company specification | Capsule shell | 1 piece |

[a] Equivalent to 50 mg free form and the amount may be adjusted based on its purity

##### 7.1.2 Reference Treatment (Placebo)

Active substance: None

**Table 5 Qualitative and Quantitative Composition of Placebo**

| Component | Quality reference | Function | Quantity (mg) /unit |
|---|---|---|---|
| Mannitol | Ph. Eur. | Filler | 347.32 |
| Anhydrous colloidal silica | Ph. Eur | Glidant | 2.94 |
| Magnesium stearate | Ph. Eur | Lubricant | 3.53 |
| Vcaps® Plus capsule | Company specification | Capsule shell | 1 piece |

##### 7.2 Packaging, Labelling and Storage

IMP will be delivered as 50 mg capsules packed in plastic containers with 28 capsules in each.

Each container will be labelled in local language.

At the trial site, IMP must be stored separately from normal clinic stocks in a securely locked area only accessible to authorised trial personnel. Storage temperature must be monitored and kept at room temperature (15-25$^{o}$ C) at the pharmacy.

Labelling of the IMP will be done in compliance with GMP Annex 13 (GMP 2003) and local regulatory requirements.

IMP will be administereded twice daily to subjects at Visits 2-8.

##### 7.3 Treatment Assignment

Eligible subjects will be randomised to receive either SoC + placebo or SoC + C21. The doses administered to each subject will be:

- C21 100 mg *b.i.d.*
- Placebo *b.i.d*

Each subject will be assigned a ransomisation (kit) code number.

Authorised staff will document the randomisation number assigned in the subject's medical record and in the eCRF.

##### 7.4 IMP Administration

IMP will be administered twice daily as follows:

- Morning dose: Two 50 mg capsules (100 mg C21 or placebo) to be taken with a glass of water after minimum 2 hours fasting
- Afternoon/evening dose: Two 50 mg capsules (100 mg C21 or placebo) to be taken with a glass of water after minimum 2 hours fasting

Subjects will be required not to eat anything for 1 hour after taking the IMP.

##### 7.5 Compliance Check and Drug Accountability

Compliance with C21 administration will be assessed by capsule count performed by the trial staff at Visit 8 or as an additional withdrawal assessment (see Section 6.5.3), if the subject is prematurely withdrawn. Details of the capsule count and compliance check will be recorded in the subject's medical record.

After IMP accountability has been completed, all unused and partly used IMP will be returned to the Sponsor or Sponsor's designee for destruction. All returned IMP will be documented and reconciled.

##### 7.6 Blinding of the Trial

The trial will be conducted in double-blind fashion and the allocation of treatments will not be disclosed until clean file has been declared and the database has been locked.

Active treatment (C21) and reference treatment (matching placebo capsules) are identical in size, colour, smell, and appearance. The packaging and labelling of IMP will reveal no evidence of IMP identity. Hence, it is expected that subjects, Investigator and other trial personnel will remain unaware of treatment allocation.

##### 7.7 Procedures for Unblinding

The Sponsor or Sponsor's designee will provide trial sites with one randomisation code envelope per each randomised subject (together with the IMP).

The treatment code may only be broken in case of a medical emergency. The Investigator shall only un-blind the treatment allocation of a subject in the course of a clinical trial, if un-blinding is relevant for the safety of the subject.

For expedited reporting purposes, the Sponsor or Sponsor's designee will be able to perform un-blinding.

Whenever a randomisation code is broken at the site, the person breaking the code must record the time, date, and reason as well as her/his initials in the subject's medical records.

The subject must be withdrawn immediately after code break.

##### 7.8 Prior and Concomitant Medications

Concomitant medications are all medications being continued by the subject at trial entry, and all medications 3 months prior to screening and received in addition to IMP during the trial period.

At each visit, the Investigator or qualified designee will record concomitant medication. All concomitant medications will be documented in the subject's medical records and in the eCRF. Any changes in concomitant medications (e.g. new treatment, discontinuation of treatment or change in dosage) during the trial period must be documented in the subject's medical records and in the eCRF.

The following information will be recorded in the eCRF:

- Generic name (preferred) or trade name
- Reason for prescription
- Dose unit and frequency
- Route of administration
- Start date (if started > 3 months prior to Visit 1, then this can be stated instead of recording a date)
- Stop date (unless ongoing at trial termination)

##### 7.8.1 Allowed Concomitant Medication

Concomitant medication may be given according to local standard of care.

##### 7.8.2 Disallowed Concomitant Medication

The following treatments are not allowed:

One week before Visit 1 and during the trial period

- Strong CYP3A4 inducers (e.g. rifampicin, phenytoin, St. John's Wort, phenobarbital, rifabutin, carbamazepine, anti HIV drugs, barbituates)
- Warfarin
- Experimental drugs

### 8 ASSESSMENTS

#### 8.1 Efficacy Assessments

##### 8.1.1 Temperature

Daily measurements of body temperature will be performed at each visit from Visit 1 (screening) to Visit 8 (end-of-treatment) in the morning.

##### 8.1.2 Laboratory Samples

The following blood samples, will be obtained at each visit from Visit 1 (screening) to Visit 8 (end-of-treatment).

- CRP (additional blood samples for CRP must be obtained from subjects prematurely withdrawn from IMP (see Section 6.5.3)
- IL-6, IL-10, TNF, CA125, and ferritin

Details of blood sampling and storage will be specified in the laboratory manual.

##### 8.1.3 Biomarkers

Blood samples for potential analysis of biomarkers related to inflammation and lung injury will be obtained at Visit 2 and Visit 8 (end-of-treatment).

Details of blood sampling and storage will be specified in the laboratory manual.

#### 8.2 Safety Assessments

##### 8.2.1 Adverse Events

Adverse events will be reported from signing of informed consent until end-of-trial participation (Visit 9).

##### 8.2.2 Medical History and Concomitant Illnesses

The medical history including date of first diagnosis of disease under study and concomitant illnesses will be obtained by interviewing the subject or by checking his/her medical records.

##### 8.2.3 Pregnancy Test

Women of childbearing potential will undergo pregnancy tests (urine dip-sticks) at Visit 1 and Visit 8 (or at the withdrawal assessments if the subject is withdrawn prematurely). If the urine pregnancy test is positive, a blood pregnancy test will be performed to confirm positive pregnancy.[1)]

1) Women of non-childbearing potential are defined as pre-menopausal females who are sterilised (tubal ligation or permanent bilateral occlusion of fallopian tubes); or post-menopausal defined as 12 months of amenorrhea).

##### 8.2.4 Physical Examination

A complete physical examination including assessments of the head, eyes, ears, nose, throat, skin, thyroid, neurological, lungs, cardiovascular, abdomen (liver and spleen), lymph nodes, and extremities will be performed at Visit 1.

A short version of the physical examination including assessments of selected body systems at the judgement of the Investigator will be performed at Visits 2-8.

##### 8.2.5 Vital Signs and Body Temperature

Systolic and diastolic blood pressure, pulse, respiration rate, and body temperature will be measured sitting in a 45$^{\circ}$ angle position after 10 minutes of rest at all visits except Visit 9 (end-of-trial).

##### 8.2.6 Electrocardiogram

12-lead electrocardiography (ECG) will be recorded at Visit 1.

##### 8.2.7 Safety Laboratory Parameters

Safety laboratory parameters which are to be taken at every visit except Visit 9 (end-of-trial):

- Haematology: haemoglobin (Hb), haematocrit (erythrocyte volume fraction), platelet count (thrombocyte particle concentration [TPC]), leucocyte count, mean corpuscular volume (MCV), white blood cells with differential count
- Biochemistry: albumin, alanine transferase (ALT), aspartate aminotransferase (AST), alkaline phosphatase (ALP), bilirubin, blood urea nitrogen (BUN), calcium, creatinine, glucose, potassium, sodium
- Urinalysis: dipstick for bilirubin, glucose, ketones, protein, specific gravity, urobilinogen, and pH

Any laboratory abnormality, judged by the Investigator to be a clinically relevant worsening since Visit 1, should be reported as an adverse event if the laboratory abnormality requires clinical intervention or further investigation, unless the laboratory abnormality is associated with an already reported event.

#### 8.3 Other Assessments

##### 8.3.1 Demographics

Demographic and baseline characteristics will be obtained at Visit 1. These include but are not limited to age at screening, sex, height, and body weight.

### 9 ADVERSE EVENTS

#### 9.1 Adverse Event Definitions

An adverse event (AE), an adverse drug reaction (ADR), and a serious adverse event (SAE) are defined according to ICH Guideline E2A (ICH 1994).

An AE is any untoward medical occurrence in a subject administered the IMP and which may or may not have a causal relationship with this IMP. An AE can therefore be any unfavorable and unintended sign (e.g. a significant abnormal laboratory finding, symptom, or disease temporally associated with the use of the IMP, whether or not considered related to the IMP.

An ADR is any noxious and unintended response to an IMP related to any dose of the IMP.

An SAE is any untoward medical occurrence that at any dose:

- Results in death
- Is life-threatening (this refers to an event in which the subject was at risk of death at the time of the event; it does not refer to an event that hypothetically might have caused death if it was more severe)
- Requires in-patient hospitalisation or prolongation of existing hospitalisation
- Results in persistent or significant disability or incapacity
- Is a congenital anomaly or birth defect

- Is judged medically important (this refers to an event that may not be immediately life-threatening or result in death or hospitalisation, but may jeopardise the subject or may require intervention to prevent one of the other outcomes listed)

A non-SAE is any AE that does not meet the definition of an SAE.

The following will not be considered an AE:

- Pre-planned procedure (documented at Visit 1) unless the condition for which the procedure was planned has worsened from the first trial related activity after the subject has signed the informed consent form
- Concomitant illness identified as a result of screening procedures. However, if symptoms are worsened and/or become serious as defined in Section 9.1, this must be reported as a SAE.

#### 9.2 Adverse Event Assessment Definitions

##### 9.2.1 Severity

The Investigator should assess the severity of all AEs according to the following definitions:

- Mild: awareness of sign or symptom, but easily tolerated (acceptable).
- Moderate: discomfort to interfere with usual activities (disturbing).
- Severe: incapacity to work or to perform usual activities (unacceptable).

Note the distinction between seriousness and severity: The term severe is used to describe the intensity of the event and a severe event is not necessarily serious. The seriousness criteria serve as a guide for defining regulatory reporting obligations.

##### 9.2.2 Relationship to IMP

Assessment of causality is based on the following considerations: associative connections (time and/or place), pharmacological explanations, previous knowledge of the drug, presence of characteristic clinical or pathological phenomena, exclusion of other causes, and/or absence of alternative explanations.

The Investigator will be asked to assess causal relationship to the IMP according to the following classifications:

*Related*:

- Time relationship exists; and previous knowledge of the trial product supports a causal relationship although another cause cannot be ruled out; and/or improvement on dechallenge or dose reduction has occurred (if performed); and/or recurrence of symptoms on rechallenge has occurred (if performed); and/or a specific laboratory test supports a causal relationship

*Not related*:

- No time relationship between administration of the trial product and occurrence or worsening of the AE exists; and/or another cause is confirmed and no indication for involvement of the trial product in the occurrence/worsening of the AE exists. The alternative, most likely other cause(s) should be indicated

##### 9.2.3 Outcome

The Investigator will be asked to record the most appropriate outcome of the following:

- Recovered/resolved
- Recovering/resolving
- Not recovered/not resolved
- Recovered/resolved with sequelae
- Fatal
- Unknown

#### 9.3 Reporting of Adverse Events

All events meeting the definition of an AE must be reported in the period from the subject has signed the informed consent form until the end-of-trial participation (Visit 9).

At each visit the subject will be asked about AEs in an objective manner, *e.g.*: "Have you experienced any problems since the last visit?"

Only medically qualified personnel (Investigators) must assess AEs.

AEs must be reported on the AE form. In the eCRF the diagnosis should be recorded, if available. If no diagnosis is available each sign and symptom should be recorded as individual AEs.

SAEs, in addition, must be reported within 24 hours of obtaining knowledge of the event. The information to be reported will include assessment of severity, causal relationship to IMP or trial procedures, action taken, outcome, and a narrative description of the course of the event. Additional information may be subsequently provided.

The SAE form and all other relevant documents supporting the reported SAE (e.g. diagnostic procedures, hospital records, autopsy reports) must be faxed or scanned/emailed to the Sponsor or Sponsor's designee.

The independent ethics committees (IEC) and regulatory authorities will be notified of SAEs according to current regulation and local requirements.

All suspected unexpected serious adverse reactions (SUSARs) are subject to expedited reporting to regulatory authorities. Pharmacovigilance will unblind such cases prior to reporting.

Investigators, the Sponsor and local IECs (if applicable) will remain blinded.

The expectedness of a SUSAR will be assessed against the reference safety information. Any SUSARs will be reported by the clinical research organisation (CRO) to the relevant competent authority. SUSARS will also be reported to the IEC by the site/CRO. Fatal and life threatening SUSARs will be reported as soon as possible and no later than seven calendar days from the date of first knowledge of the event. Relevant follow-up information will subsequently be expedited within an additional eight days. All other SUSARs will be expedited no later than 15 calendar days of first knowledge of the event.

##### 9.4 Follow-up on Adverse Events

All AEs should be followed until they are resolved or the subject's participation in the trial ends, whichever comes first.

SAEs and severe, non-serious AEs considered related to IMP should be followed on a regular basis according to the investigator's clinical judgment until a final outcome has been established.

##### 9.5 Pregnancies

Any pregnancy that occurs during trial participation must be reported and administration of IMP must be terminated immediately. A pregnancy must be reported immediately to Sponsor or designee. The pregnancy must be followed up to determine outcome (including premature termination) and status of mother and infant. Pregnancy complications and elective terminations for medical reasons must be reported as AEs or SAEs, as applicable. Spontaneous abortion must be reported as an SAE.

In addition, the Investigator must attempt to collect pregnancy information on any female partners of male trial participants who become pregnant while the participant is enrolled in the trial. Pregnancy information must be reported to Sponsor as described above.

#### 10 CHANGES TO TRIAL CONDUCT

##### 10.1 Protocol Amendments

Before implementation of substantial changes (as defined in EU guidance documents [European Commission 2006, 2010]) unless considered an Urgent Safety Measure, approval/favourable opinion must be obtained from the appropriate regulatory authority(ies) and IEC(s).

##### 10.2 Premature Termination of the Trial

In case of premature termination of the trial, regulatory authorities, and IECs will be notified in writing, including the reason for premature termination.

Conditions that may warrant premature termination of the trial include, but are not limited to the following:

- The discovery of an unexpected and significant or unacceptable risk to the subjects enrolled in the trial
- A decision of the Sponsor to discontinue development of the IMP

##### 10.3 Premature Termination of a Trial Site

The Sponsor can decide to prematurely terminate a site. Conditions that may warrant termination include, but are not limited to:

- Insufficient adherence to protocol requirements
- Failure to enrol subjects at an acceptable rate

#### 11 DATA HANDLING AND RECORD KEEPING

##### 11.1 Data from Clinical Trial Sites

Data from clinical trial sites will be entered in an eCRF.

The Investigator will sign relevant eCRF sections by use of an electronic signature. Only the Investigator (i.e. medically qualified personnel) can sign data on medical assessments. Any corrections made by the Investigator or authorised staff to the eCRF after original entry will be documented in an audit trail. The person making the change and the date, time and reason for the change will be identified in the audit trail. Changes to the data already approved/signed by an Investigator will require re-signature by the Investigator. The Investigator (principal Investigator or sub-Investigator) will sign all subject data in the eCRF by an electronic signature.

Subject data will be recorded anonymously and the subjects will be identified only by a screening number.

The monitor will check the eCRF for accuracy and completion and perform source data verification. Data entered in the eCRF will be verified against source data. The level of source data verification is described in Section 11.2.

##### 11.2 Source Data

All data entered in the eCRF should be verifiable by source data in the subject’s medical record or other records at the trial site, as applicable:

- Details of the trial (trial ID and subject screening and randomisation number)
- Date(s) of informed consent of the subject
- Data of each trial visit including signature and/or initials of person(s) conducting the visit
- Data and information of any relevant telephone contact with the subject and signature and/or initials of person(s) conducting or receiving the call
- Subject’s date of birth
- Diagnosis of coronavirus (SARS-CoV)-2 infection including start date
- Medical history and concomitant illness including start and stop dates

- Concomitant medication including start and stop dates
- Overall conclusion of the subject's eligibility
- Conclusion and results for each in-/exclusion criterion with respect to fulfillment of trial eligibility or not
- Physical examination
- Height and body weight
- Blood pressure, pulse, respiration rate, and body temperature
- ECG evaluation by Investigator
- All laboratory samplings and data, including date and time of blood and urine
- Investigator's evaluation of haematology and biochemistry results being out of range
- Date and times of blood sampling
- All AEs, SAEs, and pregnancies described in details
- Date and time of IMP administration
- Fasting criteria fulfilled
- Premature withdrawal of subject from the trial including reason and withdrawal criteria fullfilled

##### 11.3 Coding of Data

Medical history and AEs will be coded using the lastest version of Medical Dictionary for Regulatory Activities (MedDRA).

Concomitant medication will be coded using the latest version of WHO Drug Reference List.

##### 11.4 Subject Confidentiality

The confidentiality of the subject data and subject records shall be protected, respecting the privacy and confidentiality rules in accordance with applicable regulatory requirements.

#### 12 RETENTION OF DOCUMENTS

The monitor will instruct the Investigator to maintain source documents and the signed informed consent form for each subject.

Furthermore, the monitor will instruct the Investigator to archive essential documents for the duration defined in the ICH Guideline E6 (Note for Guidance on Good Clinical Practice [ICH-GCP R2]) or for 15 years, whichever comes first.

The duration of archiving defined in the ICH Guideline E6 is as follows:

Essential documents should be retained until at least 2 years after the last approval of a marketing application in an ICH region and until there are no pending or contemplated marketing applications in an ICH region or at least 2 years have elapsed since the formal discontinuation of clinical development of the investigational product. These documents should be retained for a longer period however if required by the applicable regulatory requirements or by an agreement with the Sponsor.

The Sponsor will notify the Investigator when retention of the trial-related records is no longer required.

### 13 STATISTICAL METHODS

The principal features of the statistical analysis to be performed are described in this section. A more technical and detailed elaboration of the principal features will be presented in a separate statistical analysis plan, which will be signed and approved prior to the database lock.

#### 13.1 Sample Size and Power Considerations

Sample size is based on the assumption of a standard deviation of 60 mg/L for the change in CRP. With 75 subjects per group and a two-sided t-test at 10% significance level, there will be an approximative 80% power to detect a true reduction of 25 mg/L in C21 treated subjects compared to placebo.

**Analysis Data Sets**

The full analysis set (FAS) will consist of all subjects who have been randomised and received at least one dose of IMP and who has at least one post-baseline assessment of efficacy.

The per protocol analysis set (PPAS) will be a subset of FAS and consist of all subjects without any major protocol deviations that are judged to compromise the analysis of the data. All protocol deviations will be judged as major or minor prior to database lock.

The safety analysis set (SAS) will consist of all subjects who have been randomised and received at least one dose of IMP.

#### 13.2 Subject Disposition

##### 13.2.1 Baseline Characteristics

The subject flow in terms of number of enrolled, randomised, treated, completed, and withdrawn (including reason for withdrawal) subjects and subjects included in each analysis data set will be summarised by treatment group.

Demographics, medical history, baseline characteristics, and prior medications will be listed and summarised by treatment group in terms of descriptive statistics: mean, standard deviation, median, and range for continuous outcomes and frequencies and percentages for categorical outcomes.

#### 13.3 Efficacy Analysis

##### 13.3.1 Efficacy Endpoint Analysis

The primary endpoint will be the change in CRP from baseline to the average of the last 2 assessments during the treatment period. For early withdrawn subjects (any cause) the last computable average or value (if only one) will be carried forward to represent state at end-of-trial. CRP data will be summarised by day and treatment including the change from baseline. Treatments will be compared using an analysis of covariance (ANCOVA) model with fixed factor treatment and baseline CRP as covariate. Estimated treatment difference will be given with 90% confidence intervals and associated, two-sided, p-value. Subjects will be evaluated both with respect to the FAS and the PPAS populations.

Secondary (continuous) endpoints such as body temperature and pre-specified laboratory values and biomarkers will be summarised by visit and for the change over trial by descriptive statistics. The change from baseline will be compared using similar ANCOVA models as the primary endpoint.

The number of subjects not in need of oxygen or not in need of mechanical ventilaton will be compared between treatments using a logistic regression model adjusting for treatment. Estimated treatment difference will be given as an odds ratio with 90% confidence intervals and associated, two-sided, p-value. For early withdrawn subjects the status as assessed on last day in the treatment will be used.

The time to need of mechanical invasive or non-invasive ventilation will be summarised by treatment group and compared between groups using the log-rank test. Subjects not in need of mechanical ventilation will be censored at last day in treatment period.

The time on oxygen supply (for those not needing mechanical ventilation) will be summarised by treatment group. For subjects withdrawing from the trial (for instance due to worsening), imputation assuming oxygen need will be made. Treatment groups will be compared using the Wilcoxon rank sum test.

##### 13.3.2 Biomarker Data

Biomarker data will be exploratory and the analyses data-driven. Collected data will be visualised graphically and summarised descriptively.

##### 13.3.3 Statistical/Analytical Issues

###### *13.3.3.1 Missing Data*

The imputation of last value (average) carried will be used for analysis if subject is early withdrawn.

###### *13.3.3.2 Examination of Subgroups*

Important demographic and baseline value-defined subgroups may be investigated. A detailed description will be provided in the Statistical Analysis Plan.

#### 13.4 Safety Analysis

##### 13.4.1 Adverse Events

An overview of all treatment-emerging AEs including severity, relationship to IMP, SAEs and AEs leading to withdrawals of treatment and withdrawal from trial or death will be presented by treatment group. All treatment-emergent AEs will be listed.

AEs will be summarised by system organ class (according to MedDRA), and preferred term (according to MedDRA) displaying number of subjects in each treatment group, number and percentage of subjects having the AE as well as number of AEs. Furthermore, AEs will be summarised according to severity and relationship to IMP.

SAEs and AEs leading to withdrawal from treatment will be listed and tabulated by System Organ Class and preferred term, if appropriate.

##### 13.4.2 Vital Signs

Vital signs data (e.g. blood pressure and body temperature) will be summarised by treatment group for each visit including the change over trial using descriptive statistics.

##### 13.4.3 Laboratory Safety Assessments

Laboratory test data will be summarised by treatment group and visit, and for changes over trial using descriptive statistics. Abnormal laboratory test results will be summarised by treatment and visit using numbers and frequencies.

##### 13.4.4 Physical examination

Abnormal findings on the physical examination will be summarised by visit.

### 14 INDEPENDENT DATA MONITORING COMMITTEE

Not applicable.

### 15 GOOD CLINICAL PRACTICE

This trial will be carried out in compliance with the protocol, ICH-GCP R2, standard operating procedures and applicable regulatory requirements.

The Investigator agrees, when signing this protocol, to adhere to the instructions and procedures described in it, to the principles of the Declaration of Helsinki, ICH-GCP R2, and applicable regulatory requirements.

### 16 ETHICS

#### 16.1 Independent Ethics Committees / Health Authorities

Before implementing this trial, the protocol, the proposed subject information and subject consent form, and other documents as required, will be reviewed by properly constituted IECs and by the national regulatory authorities.

A signed and dated statement that the protocol and subject information and subject consent form have been approved by the IECs and regulatory authorities will be obtained before trial initiation.

For each individual IEC, the name and occupation of the chairman and the members of the IEC will be collected as well as a statement that the IEC works in accordance with ICH GCP.

IECs will receive updates on trial progress according to local regulations.

#### 16.2 Informed Consent

The subject's signed and dated informed consent to participate in the trial will be obtained prior to any trial-related procedures being carried out.

Before any trial-related procedures, the investigator will explain to the potential subject the aims, methods, reasonably expected benefits, and potential hazards of the trial and any discomfort participation in the trial may entail. Subjects will be informed that participation in the trial is voluntary and that the subject may withdraw from the trial at any time and for any reason. Subjects will be informed that if they choose not to participate, this will not affect the care the subject will receive for the treatment of his or her disease. Finally, subjects will be informed that their records may be accessed by health authorities and authorised Sponsor staff without violating the confidentiality of the subject, to the extent permitted by the applicable laws or regulations.

All subjects will be given opportunity to ask questions and will be given sufficient time to consider before consenting. The subjects may choose to be accompanied, e.g. by a family member, during the information process. After having consented, a copy of the informed consent form will be given to the subject.

### 17 AUDITS AND INSPECTIONS

A representative of the Sponsor or the Sponsor may visit the trial site(s) at any time during the trial or after completion of the trial to conduct an audit of the trial. These audits will require access to all trial records, including source documents, for inspection and comparison with the CRFs. Subject privacy will, however, be respected. The Investigator and other trial personnel will be responsible for being present and available for consultation during routinely scheduled site audit visits conducted by the Sponsor's representative.

Similar auditing procedures (inspections) may also be conducted by agents of regulatory health authorities, either as part of a national GCP compliance program or to review the results of this trial in support of a regulatory submission. The Investigator should notify the Sponsor's representative or Sponsor immediately, if he/she has been contacted by a regulatory agency concerning an upcoming inspection.

### 18 MONITORING

Before trial initiation a monitor from the Sponsor's representative will review the protocol and the CRF with the Investigators and their trial personnel. During the trial the monitor will check the completeness of subject records, the accuracy of entries in CRFs, the adherence to the protocol and to GCP, the progress of enrolment and the handling and accounting of IMP. Key trial personnel must be available to support the monitor.

The Investigator must give the monitor direct access to source data/documents (e.g. relevant hospital or medical records) to confirm their consistency with the entries in CRFs. The Sponsor representative will ensure remote access to records will be by authorised personnel only.

### 19 REPORTING OF RESULTS

#### 19.1 Integrated Clinical Trial Report

Data will be reported in an integrated clinical trial report in compliance with the requirements of the current version of ICH E3: Structure and Content of Clinical Study Report (ICH 1995). The signatory Investigator will review and sign the integrated clinical trial report.

The results and interpretation of the exploratory biomarker analyses may not be included in the clinical trial report but may be reported and/or published separately at a later stage.

#### 19.2 Use of Information

All unpublished information relating to this trial and/or to the IMP is considered confidential by the Sponsor and shall remain the sole property of the Sponsor.

The Investigator must accept that the Sponsor may use the information from this clinical trial in connection with the development of the IMP, and therefore, may disclose it as required to other investigators, to government licensing authorities, to regulatory agencies of other governments, stock exchange market, and commercial partners.

##### 19.3 Publication of Results

The Sponsor is committed to publishing the trial results, whether positive or negative, in a peer-reviewed journal (Wager, 2003; Graf 2009).

The criteria for authorship as set out by the Committee of Medical Journal Editors (www.icmje.org) will be applied.

The contributorship model will be applied and contributors who do not meet the criteria for authorship will be listed in an acknowledgments section with descriptions of the role of each contributor in order to ensure indexation in the National Library of Medicine.

Publications are subject to the following conditions:

- Data are the property of the Sponsor and cannot be published without prior authorisation from the Sponsor
- Publications should be drafted with protection of individual privacy, intellectual property, and contract rights in mind, and also conform to legislation and current national practices in patent and other laws
- The primary publication (i.e. the results from all centers) should be published before, or in parallel with, any secondary publications
- Publications shall not disclose any Sponsor confidential information or property

#### 20 INSURANCE AND LIABILITY

The Sponsor has subscribed to an insurance policy covering, in its terms and provision, its legal liability for injuries caused to participating subjects and arising out of trial procedures performed in accordance this protocol, in accordance with applicable law and with the ICH Guideline E6 (Note for Guidance on Good Clinical Practice) (ICH-GCP R2).

European Commission 2010 Detailed guidance for the request for authorization of a clinical trial on a medicinal product for human use to the competent authorities, notification of substantial amendments and declaration of the end of the trial (CT1).

European Commission 2006 Detailed guidance on the application format and documentation to be submitted in an application for an Ethics Committee opinion on the clinical trial on medicinal products for human use (CT2).

Graf, C., Battista, W. P., Bridges, D., Bruce-Winkler, V., Conaty, J. M., Ellison, J. M., Field, E. A., Gurr, J. A., Marx, M. E., Patel, M., Sanes-Miller, C., Yarker, Y. E. International Society for Medical Publication Professional. 2009 Good publication practise for community company Sponsored medical research: the GPP2 guidelines. BMJ 339:b4330.

Good Manufacturing Practices. 2003 Annex 13. Manufacture of Investigational medicinal products.

Guignabert, C., de Man, F., Lombès, M. 2018 ACE2 as therapy for pulmonary arterial hypertension: the good outweighs the bad. Eur. Respir. J. 51(6).

Hanff, T. C., Harhay, M. O., Brown, T. S., Cohen, J. B., Mohareb, A. M. 2020 Is there an association between COVID-19 mortality and the renin-angiotensin system – A call for epidemiologic investigations. Clin Infect Dis. 2020 Mar 26. pii: ciaa329. doi: 10.1093/cid/ciaa329. Epub ahead of print.

Henderson, L. A., Canna, S. W., Schulert, G. S., Volpi, S., Lee, P. Y., Kernan, K. F., Caricchio, R., Mahmud, S., Hazen, M. M., Halyabar, O., Hoyt, K. J., Han, J., Grom, A. A., Gattorno, M., Ravelli, A., de Benedetti, F., Behrens, E. M., Cron, R. Q., Nigrovic, P. A. 2020 On the alert for cytokine storm: Immunopathology in COVID-19. Arthritis Rheumatol. 2020 Apr 15. doi: 10.1002/art.41285. [Epub ahead of print]

Huang, C., Wang, Y., Li, X., Ren, L., Zhao, J., Hu, Y., Zhang, L., Fan, G., Xu, J., Gu, X., Cheng, Z., Yu, T., Xia, J., Wei, Y., Wu, W., Xie, X., Yin, W., Li, H., Liu, M., Xiao, Y., Gao, H.,

Imai, Y., Kuba, K., Rao, S., Huan, Y., Guo, F., Guan, B., Yang, P., Sarao, R., Wada, T., Leong-Poi, H., Crackower, M. A., Fukamizu, A., Hui, C. C., Hein, L., Uhlig, S., Slutsky, A. S., Jiang, C., Penninger, J. M. 2005 Angiotensin-converting enzyme 2 protects from severe acute lung failure. Nature 436(7047):112-6.

Guo, L., Xie, J., Wang, G., Jiang, R., Gao, Z., Jin, Q., Wang, J., Cao, B. (2020) Clinical features of patients infected with 2019 novel coronavirus in Wuhan, China. Lancet 395(10223):497-506.

Human Protein Atlas, https://www.proteinatlas.org/ENSG00000180772-AGTR2

ICH Harmonized Tripartite Guideline E3. 1995 Structure and Content of Clinical Study Reports. Step 5.

ICH Harmonized Tripartite Guideline E2A. 1994 Step 5: Clinical Safety Data Management: Definitions and Standards for Expedited Reporting.

ICH Harmonized Tripartite Guideline E6 (R2). 1997 Step 5: Guideline for Good Clinical Practice. Adopted by CPMP Dec 2016.

ICNARC 2020. Case Mix Programme Database 17 April 2020: ICNARC report on COVID-19 in critical care.

Kuba, K., Imai, Y., Rao, S., Gao, H., Guo, F., Guan, B., Huan, Y., Yang, P., Zhang, Y., Deng, W., Bao, L., Zhang, B., Liu, G., Wang, Z., Chappell, M., Liu, Y., Zheng, D., Leibbrandt, A., Wada, T., Slutsky, A. S., Liu, D., Qin, C., Jiang, C., Penninger, J. M. (2005) A crucial role of angiotensin converting enzyme 2 (ACE2) in SARS coronavirus-induced lung injury. Nat. Med. 11(8):875-9.

Lake, M. A. 2020 What we know so far: COVID-19 current clinical knowledge and research. Clinical Medicine 20(2):124–127.

Li, Q., Guan, X., Wu, P., Wang, X., Zhou, L., Tong, Y., Ren, R., Leung, K. S. M., Lau, E. H. Y., Wong, J. Y., Xing, X., Xiang, N., Wu, Y., Li, C., Chen, Q., Li, D., Liu, T., Zhao, J., Liu, M., Tu, W., Chen, C., Jin, L., Yang, R., Wang, Q., Zhou, S., Wang, R., Liu, H., Luo, Y., Liu, Y., Shao, G., Li, H., Tao, Z., Yang, Y., Deng, Z., Liu, B., Ma, Z., Zhang, Y., Shi, G., Lam, T. T. Y., Wu, J. T., Gao, G. F., Cowling, B. J., Yang, B., Leung, G. M., Feng, Z. (2020) Early Transmission Dynamics in Wuhan, China, of Novel Coronavirus–Infected Pneumonia. N. Engl. J. Med. 382(13):1199-1207.

Li, W., Moore, M. J., Vasilieva, N., Sui, J., Wong, S. K., Berne, M. A., Somasundaran, M., Sullivan, J. L., Luzuriaga, K., Greenough, T. C., Choe, H., Farzan, M. (2003) Angiotensin-converting enzyme 2 is a functional receptor for the SARS coronavirus. Nature 426(6965):450-454.

Lu, R., Zhao, X., Li, J., Niu, P., Yang, B., Wu, H., Wang, W., Song, H., Huang, B., Zhu, N., Bi, Y., Ma, X., Zhan, F., Wang, L., Hu, T., Zhou, H., Hu, Z., Zhou, W., Zhao, L., Chen, J., Meng, Y., Wang, J., Lin, Y., Yuan, J., Xie, Z., Ma, J., Liu, W. J., Wang, D., Xu, W., Holmes, E. C., Gao, G. F., Wu, G., Chen, W., Shi, W., Tan, W. (2020) Genomic characterisation and epidemiology of 2019 novel coronavirus: implications for virus origins and receptor binding. Lancet 395(10224):565-574.

Matavelli, L. C. and Siragy, H. M. 2015 AT2 receptor activities and pathophysiological implications. J. Cardiovasc. Pharmacol. 65(3):226-32.

Menk, M., Graw, J. A., von Haefen, C., Steinkraus, H., Lachmann, B., Spies, C. D., Schwaiberger, D. (2018) Angiotensin II type 2 receptor agonist Compound 21 attenuates pulmonary inflammation in a model of acute lung injury. J. Inflamm. Res. 11:169-178.

Phan, L. T., Nguyen, T. V., Luong, Q. C., Nguyen, T. V., Nguyen, H. T., Le, H. Q., Nguyen, T. T., Cao, T. M., Pham, Q. D. (2020) Importation and Human-to-Human Transmission of a Novel Coronavirus in Vietnam. N. Engl. J. Med. 382(9):872-874.

Rathinasabapathy, A., Horowitz, A., Horton, K., Kumar, A., Gladson, S., Unger, T., Martinez, D., Bedse, G., West, J., Raizada, M.K., Steckelings, U.M., Sumners, C., Katovich, M.J., Shenoy, V. 2018 The selective angiotensin II type 2 receptor agonist compound 21, attenuates the progression of lung fibrosis and pulmonary hypertension in an experimental model of bleomycin-induced lung injury Front. Physiol. 9:180.

Ruan, Q., Yang, K., Wang, W., Jiang, L., Song, J. (2020) Clinical predictors of mortality due to COVID-19 based on an analysis of data of 150 patients from Wuhan. China Intensive Care Med. Epub ahead of print. https://doi.org/10.1007/s00134-020-05991-x.

Steckelings, U. M., Kaschina, E., Unger, Th. 2005 The AT2 receptor - A matter of love and hate. Peptides 26:1401-1409.
Tan, L., Wang, Q., Zhang, D., Ding, J., Huang, Q., Tang, Y. Q., Wang, Q., Miao, H. (2020) Lymphopenia predicts disease severity of COVID-19: a descriptive and predictive study. Signal Transduct. Target Ther. 5(1):33.

Verity, R., Okell, L. C., Dorigatti, I., Winskill, P., Whittaker, C., Imai, N., Cuomo-Dannenburg, G., Thompson, H., Walker, P. G. T., Fu, H., Dighe, A., Griffin, J. T., Baguelin, M., Bhatia, S., Boonyasiri, A., Cori, A., Cucunubá, Z., FitzJohn, R., Gaythorpe, K., Green, W., Hamlet, A., Hinsley, W., Laydon, D., Nedjati-Gilani, G., Riley, S., van Elsland, S., Volz, E., Wang, H., Wang, Y., Xi, X., Donnelly, C. A., Ghani, A. C., Ferguson, N. M. (2020) Estimates of the severity of coronavirus disease 2019: a model-based analysis. Lancet Infect. Dis. Epub ahead of print. https://doi.org/10.1016/ S1473-3099(20)30243-7.

Wager, E., Field, E. A., Grossman, L. 2003 Good publication practices for pharmaceutical companies: why we need another set of guidelines. Curr. Med. Res. Opin. 19(3):147-8.

Wang, W., Tang, J., Wei, F. (2020) Updated understanding of the outbreak of 2019 novel coronavirus (2019-nCoV) in Wuhan, China. 2020. Journal of Medical Virology.

WHO: https://www.who.int/emergencies/diseases/novel-coronavirus-2019

WHO (2020) A coordinated global research roadmap: 2019 novel coronavairus.

Wu, F., Zhao, S., Yu, B., Chen, Y. M., Wang, W., Song, Z. G., Hu, Y., Tao, Z. W., Tian, J. H., Pei, Y. Y., Yuan, M. L., Zhang, Y. L., Dai, F. H., Liu, Y., Wang, Q. M., Zheng, J. J., Xu, L., Holmes, E. C., Zhang, Y. Z. 2020 A new coronavirus associated with human respiratory disease in China. Nature 579:265–269

Wu, P., Hao, X., Lau, E. H. Y., Wong, J. Y., Leung, K. S. M., Wu, J. T., Cowling, B. J., Leung, G. M. (2020) Real-time tentative assessment of the epidemiological characteristics of novel coronavirus infections in Wuhan, China, as at 22 January 2020. Euro Surveill. 25(3).

Zhang, H., Penninger, J. M., Li, Y., Zhong, N., Slutsky, A. S. 2020 Angiotensin-converting enzyme 2 (ACE2) as a SARS-CoV-2 receptor: molecular mechanisms and potential therapeutic target. Intensive Care Med. 46(4):586-590.

Zhou, P., Yang, X. L., Wang, X. G., Hu, B., Zhang, L., Zhang, W., Si, HR., Zhu, Y., Li, B., Huang, C. L., Chen, H. D., Chen, J., Luo, Y., Guo, H., Jiang, R. D., Liu, M. Q., Chen, Y., Shen, X. R., Wang, X., Zheng, X. S., Zhao, K., Chen, Q. J., Deng, F., Liu, L. L., Yan, B., Zhan, F. X., Wang, Y. Y., Xiao, G. F., Shi, Z. L. (2020) A pneumonia outbreak associated with a new coronavirus of probable bat origin. Nature. 579(7798):270-273.

### 22 ABBREVIATIONS

| | |
|---|---|
| Ab | Antibody |
| ACE | Aniotensin-converting enzyme |
| ADR | Adverse drug reaction |
| AE | Adverse event |
| Ag | Antigen |
| ALP | Alkaline phosphatase |
| ALT | Alanine aminotransferase |
| ANCOVA | Analysis of covariance |
| AngI | Angiotensin I |
| AngII | Angiotensin II |
| ARDS | Acute respiratory distress syndrome |
| AST | Aspartate aminotransferase |
| $AT_1R$ | Angiotension type 1 receptor |
| $AT_2R$ | Angiotension type 2 receptor |
| *b.i.d.* | bis in die (i.e. twice a day) |
| BMI | Body mass index |
| COPD | Chronic obstructiv epulmonary disease |
| CoV | Coronavirus |
| COVID | Coronavirus disease |
| CRF | Case report form |
| CRO | Contract research organisation |
| CRP | C-reactive protein |
| CYP | Cytochrome p450 |
| eCRF | Electronic case report form |
| ECG | Electrocardiography |
| FAS | Full analysis set |
| GCP | Good Clinical Practice |
| GMP | Good Manufacturing Practice |
| Hb | Haemoglobin |
| HBsAg | Hepatitis B surface antigen |
| HCVAb | Hepatitis C virus antibody |
| HIV | Human immunodeficiency virus |
| ICH | International Conference on Harmonisation |
| IEC | Independent ethics committee |
| IL | Interleukin |
| IMP | Investigational medicinal product |
| IPF | Idiopathic pulmonary fibrosis |
| MAD | Multiple ascending dose |
| MCV | Mean corpuscular volume |
| MedDRA | Medical Dictionary for Regulatory Activities |
| NOAEL | No observed adverse effect level |
| PCR | Polymerase chain reaction |
| PPAS | Per protocol analysis set |
| $R_0$ | Basic reproduction number |
| RAS | Renin-angiotensin system |
| RP | Raynaud’s phenomenon |
| SAD | Single ascending dose |
| SAE | Serious adverse event |
| SAS | Safety analysis set |
| SoC | Standard of Care |

| | |
|---|---|
| SSc | Systemic sclerosis |
| SSc-RP | Raynaud’s phenomenon secondary to SSc |
| SUSAR | Suspected unexpected serious adverse reaction |
| TEAE | Treatment-emergent adverse event |
| $t_{max}$ | Maximal plasma concentration |
| TNF | Tumor necrosis factor |
| TPC | Thrombocyte particle concentration |
| WHO | World Health Organisation |

### VP-C21-006-CTP-v2.0-20200814

Final Audit Report 2020-08-23

| | |
|---|---|
| Created: | 2020-08-14 |
| By: | Kamilla Overbeck Nygaard |
| Status: | Signed |
| Transaction ID: | CBJCHBCAABAAjkkksi5QbGBuBhDPQixDB-AYkRWWd8ab |

#### "VP-C21-006-CTP-v2.0-20200814" History

Document created by Kamilla Overbeck Nygaard
2020-08-14 - 11:06:00 AM GMT- IP address: 2.130.174.80

2020-08-14 - 11:10:20 AM GMT

2020-08-14 - 12:21:15 PM GMT- IP address: 77.215.239.209

Document e-signed by Mimi Flensburg
Signature Date: 2020-08-14 - 12:22:11 PM GMT - Time Source: server- IP address: 77.215.239.209

2020-08-14 - 12:22:13 PM GMT

2020-08-14 - 1:10:18 PM GMT- IP address: 81.141.62.76

Document e-signed by Rohit Batta
Signature Date: 2020-08-14 - 1:18:45 PM GMT - Time Source: server- IP address: 81.141.62.76

2020-08-14 - 1:18:47 PM GMT

2020-08-14 - 2:30:27 PM GMT- IP address: 98.128.181.79

Document e-signed by Thomas Bengtsson
Signature Date: 2020-08-14 - 2:31:25 PM GMT - Time Source: server- IP address: 98.128.181.79

2020-08-14 - 2:31:27 PM GMT

2020-08-23 - 8:57:57 PM GMT- IP address: 104.47.2.254

Document e-signed by Joanna C Porter

Signature Date: 2020-08-23 - 8:59:20 PM GMT - Time Source: server- IP address: 86.151.35.133

Signed document emailed to Rohit Batta, Thomas Bengtsson, Kamilla Overbeck Nygaard, Joanna C Porter, and 1 more

2020-08-23 - 8:59:20 PM GMT

Adobe Sign
