## Supplementary material for "The angiotensin type 2 receptor agonist C21 restores respiratory function in COVID19 - a double-blind, randomized, placebo-controlled Phase 2 trial": Statistical Analysis Plan

### A Randomised, Double-Blind, Placebo-Controlled, Phase 2 Trial Investigating the Safety and Efficacy of C21 in Hospitalised Subjects with COVID-19 Infection Not Requiring Mechanical Ventilation

**Vicore Pharma AB Study No: VP-C21-006**
**Syne qua non Ltd Study No: OCH20005**

### Statistical Analysis Plan

**Version: Final 1.0**

**Date: 10 September 2020**

#### Author

SignNow e-signature ID: 411ef847d0...
09/10/2020 13:23:09 UTC
Kayleigh Wolfe (Signer)

Lead Statistician

#### Approved By Orphan Reach Limited

*If signing manually, please include: Signature + Date + Full Name + Position*

SignNow e-signature ID: 2f9727c9eb...
09/10/2020 13:39:20 UTC
Gajendrasinh Chanchu (Signer)

#### LIST OF ABBREVIATIONS

| | |
|---|---|
| AE | Adverse Event |
| ANCOVA | Analysis of Covariance |
| b.i.d | *bis in die* |
| BMI | Body Mass Index |
| CI | Confidence Interval |
| CRP | C-reactive Protein |
| CV | Coefficient of Variation |
| ECG | Electrocardiogram |
| eCRF | Electronic Case Report Form |
| FAS | Full Analysis Set |
| IMP | Investigational Medicinal Product |
| KM | Kaplan Meier |
| LS Means | Least Square Means |
| MedDRA | Medical Dictionary for Regulatory Activities |
| PDC | Protocol Deviation Criteria |
| PPAS | Per-Protocol Analysis Set |
| PT | Preferred Term |
| RAS | Randomised Analysis Set |
| SAP | Statistical Analysis Plan |
| SAS | Safety Analysis Set |
| SD | Standard Deviation |
| SE | Standard Errors |
| SOC | System Organ Class |
| SoC | Standard of Care |
| TEAE | Treatment-Emergent Adverse Event |
| WHO Drug | World Health Organization Drug Dictionary |

### 1 INTRODUCTION

This document details the statistical analysis of the data that will be performed for the Vicore Pharma AB study: A randomised, double-blind, placebo-controlled, Phase 2 trial investigating the safety and efficacy of C21 in hospitalised subjects with COVID-19 infection not requiring mechanical ventilation.

The proposed analysis is based on the contents of the Final Version 1.0 of the protocol (dated 24 April 2020) and Final Version 2.0 of the protocol (dated 14 August 2020). In the event of future amendments to the protocol, this statistical analysis plan (SAP) may be modified to account for changes relevant to the statistical analysis.

The table, listing and figure shells are supplied in a separate document.

### 2 STUDY OBJECTIVES AND DESIGN

#### 2.1 Study Objectives

##### 2.1.1 Primary Objective

The primary objective of the study is to investigate the efficacy of C21 200 mg daily dose (100 mg *b.i.d.*) on COVID-19 infection not requiring mechanical invasive or non-invasive ventilation.

##### 2.1.2 Secondary Objectives

The secondary objectives of the study are to evaluate the following of C21 200 mg daily dose (100 mg *b.i.d.*):

- Effect on inflammation,
- Safety profile.

##### 2.1.3 Exploratory Objective

To investigate a range of laboratory parameters as potential biomarkers of inflammation and viral load, following oral administration of C21 200 mg daily dose (100 mg *b.i.d.*).

#### 2.2 Study Endpoints

##### 2.2.1 Primary Endpoint

The primary endpoint of the study is the change from baseline in C-reactive protein (CRP) after treatment with C21 200 mg daily dose (100 mg *b.i.d.*).

##### 2.2.2 Secondary Endpoints

The secondary endpoints are:

- Change from baseline in:
    - Body temperature,
    - IL-6,
    - IL-10,
    - TNF,
    - CA125,

    - Ferritin.
- Number of subjects not in need of oxygen supply.
- Number of subjects not in need of mechanical invasive or non-invasive ventilation.
- Time to need of mechanical invasive or non-invasive ventilation.
- Time on oxygen supply*.
- Adverse events (AEs).

* This endpoint has been amended to remove reference to those not needing mechanical invasive or non-invasive ventilation. Further information can be found in Section 6.12 of this SAP.

##### 2.2.3 Exploratory Endpoints

Blood samples will be stored for potential future analyses of biomarkers reflecting inflammation and lung injury.

#### 2.3 Study Design

The trial will be conducted at trial sites within the UK (1 site) and India (up to 12 sites). Approximately 150 subjects will be enrolled and randomised to receive twice daily oral administration of either standard of care (SoC) + placebo (N=75) or SoC + C21 (N=75). Subjects will be treated for 7 days. All subjects will be followed up 7-10 days after receiving the last investigational medicinal product (IMP) dose (visit, or phone call if recovering at home).

#### 2.4 Visit Structure

A total of 9 visits are defined:

- Visit 1: Screening,
- Visits 2–8: Treatment period,
- Visit 9: End-of-trial.

The maximum duration of the trial for any subject will be approximately 3 weeks, including a screening period of up to 3 days, a treatment period of 7 days and a visit for safety follow-up (end of-trial) to be conducted 7-10 days after the last dose of IMP.

The visit structure and scheduled assessments are detailed in Section 6, Tables 1 and 2 and Figure 1 of the protocol.

### 3 SAMPLE SIZE

Sample size is based on the assumption of a standard deviation (SD) of 60 mg/L for the change in CRP from baseline to average of last two assessments during one week of treatment. With 75 patients per group and a two-sided t-test at 10% significance level, there will be an approximative 80% power to detect a true reduction of 25 mg/L in C21 treated patients compared to placebo.

### 4 RANDOMISATION

Subjects will be randomised in a 1:1 schedule to receive either SoC + placebo or SoC + C21. Each subject will be assigned a randomisation (kit) number according to the randomisation schedule. Details of the randomisation including date of randomisation, assigned kit number, allocated treatment and treatment actually received will be listed.

#### 4.1 Planned Treatment and Actual Treatment Received

Planned treatment is defined as the treatment to which the subject was randomised and will be determined by the original randomisation list. Actual treatment received is defined as the actual study treatment received regardless of randomised treatment.

Planned study treatment will be determined from the randomisation list supplied by Vicore Pharma AB as compiled by Ardena. Actual treatment received will be derived by reconciling the kit list number entered into the IMP Administration Electronic Case Report Form (eCRF) pages with the kit list also supplied by Vicore Pharma AB as compiled by Ardena. The kit list number will be entered into the eCRF at each dose of treatment. Subjects who were mis-randomised or received treatment incorrectly will be confirmed once the blind has been broken.

### 5 INTERIM ANALYSIS

No interim analysis will be performed.

### 6 ANALYSIS PLAN

#### 6.1 General

Summary statistics for continuous variables will consist of number of non-missing observations (n), mean, SD, minimum, median and maximum, unless specified otherwise.

For categorical variables the number and percentage of subjects in each category will be presented, based on the number of non-missing observations apart from disposition of subjects, protocol deviations, background and demographic characteristics, prior and concomitant medications/procedures and AEs where the percentage will be based on the number of subjects in the analysis set with a missing category presented where applicable.

The mean and median will be displayed to 1 more decimal place than the original data values and the SD will be displayed to 2 more decimal places than the original data values. The minimum and maximum will be displayed to the same number of decimal places as the original data. Percentages will be displayed to 1 decimal place; except percentages will not be presented when the count is zero and 100% for count data will be presented as an integer.

All statistical tests will be performed using a two-tailed 10% overall significance level, unless otherwise stated. The null hypothesis at all times will be that the treatments are equivalent. All comparisons between the treatments will be reported with 90% confidence intervals (CIs) for the difference.

#### 6.2 General Derivations

This section provides details of general derivations. Derivations specific to the parameter of interest are detailed within the specific SAP section.

##### 6.2.1 Definition of Baseline

The derivation of baseline for CRP is described in Section 6.10.1 of this SAP.

Baseline for all other variables is defined as the last non-missing value (including scheduled and unscheduled assessments) prior to the subject receiving study treatment. For assessments performed at Visit 2 where it is not clear that the assessment was performed after study treatment administration, the assessment will be taken as a post-baseline assessment.

##### 6.2.2 Visit and Study Day

Throughout this SAP any references to “Visit XX” refer to the pre-specified visits defined in the protocol (where the study treatment administration visit is Visit 2).

Data listings by visit will additionally present study day, indicating the duration of time since first study treatment. Study day will be calculated relative to the first date of administration of study treatment (Study Day 1).

##### 6.2.3 Incomplete Dates

For calculation purposes, incomplete dates will be completed using worst case. Further details are given in the relevant sections as required.

##### 6.2.4 Non-numeric Values

For clinical laboratory parameters and cytokine data, in the case where a variable is recorded as “>x”, “≥x”, “<x” or “≤x” rather than an exact number, then for analysis purposes a value of x will be taken. Where a range of values is quoted the midpoint of the range will be taken. Where this process is not appropriate, further information will be detailed in the relevant section. The recorded value will be reported in listings.

##### 6.2.5 Methods for Handling Withdrawals and Missing Data

Where imputation may be deemed appropriate, information is detailed in the analysis descriptions.

#### 6.3 Analysis Sets

The **Enrolled Set** includes all subjects who provide informed consent irrespective of whether they received the study treatment.

The **Randomised Analysis Set** (RAS) includes all subjects who have been randomised, irrespective of whether they received the study treatment.

The **Full Analysis Set** (FAS) will consist of all subjects who have been randomised and received at least one dose of IMP and who have at least one post-baseline assessment of efficacy. A post-baseline assessment of efficacy includes an assessment of any of the parameters detailed in Section 6.10 of this SAP. Subjects will be analysed according to the treatment they are assigned to at randomisation, irrespective of what treatment they actually received.

The **Per-Protocol Analysis Set** (PPAS) will be a subset of the FAS and consist of all subjects without any major protocol deviations that are judged to compromise the analysis of the data. All protocol deviations will be judged as major or minor prior to database lock.

The **Safety Analysis Set** (SAS) consists of all subjects who have received at least one dose of IMP. Subjects will be analysed according to the treatment actually taken.

The list of subjects included in the PPAS will be agreed prior to breaking the blind once all study data is available. The definitions for the enrolled set, RAS, FAS and SAS are sufficient to determine the subjects included within these analysis sets and so do not require listing and agreeing prior to breaking the blind.

#### 6.4 Data Presentations

The data will be summarised in tabular form by treatment group apart from disposition of subjects, prior and concomitant medications and background and demographic data which will be summarised by treatment group and overall subjects.

Treatment groups will be presented in the following order using these labels:

- SoC + C21 100 mg b.i.d
- SoC + Placebo

Listings will be sorted by treatment group, site and subject number and date/time of assessment.

Data will be summarised and listed using the following analysis sets:

| | Enrolled | RAS | FAS | PPAS | SAS |
|---|---|---|---|---|---|
| Tables | • Disposition | • Withdrawal | • Demography[1]<br>• Efficacy | • Demography[1]<br>• Efficacy (primary endpoint only) | • Demography[1]<br>• Prior and concomitant medications<br>• Medical history<br>• Exposure<br>• Safety |
| Listings | • Completion/ withdrawal<br>• Eligibility | • Protocol deviations<br>• Analysis sets<br>• Demography<br>• Medical history<br>• Disease under investigation<br>• Hospitalisation<br>• Visit dates<br>• Randomisation<br>• Efficacy<br>• Biomarkers | | | • Prior and concomitant medications<br>• Safety<br>• IMP administration<br>• Exposure |

1. Demography will be summarised using the FAS and repeated for the PPAS and SAS where the analysis sets differ for at least 1 subject. If either of these analysis sets are identical to the FAS the summary will not be presented.

Only scheduled post baseline data will be tabulated, post-baseline repeat/unscheduled assessments will not be included in tables, although they will be listed and in particular all clinically significant values will be noted.

#### 6.5 Disposition of Subjects

The number and percentage of all subjects enrolled, included in the RAS, FAS, PPAS and SAS, who completed study treatment and prematurely discontinued study treatment, who completed the study and prematurely discontinued the study and study duration will be summarised. The number and percentage of subjects will be summarised by their reasons for withdrawal from treatment and the study.

Study duration will be derived as the number of days between date of randomisation and the date of study completion or the date of early study withdrawal. The timing of withdrawals will be explored using the RAS. Time to withdrawal from study treatment will be calculated in hours as (Date and time of withdrawal from treatment) – (Date and time of informed consent). For subjects not withdrawing from treatment, time to withdrawal will be censored at the time of their last study treatment administration. The time to withdrawal will also be presented graphically using a Kaplan Meier (KM) plot. KM estimates for the $25^{th}$, $50^{th}$ and $75^{th}$ percentiles of time to withdrawal will also be presented.

Eligibility for each of the analysis sets along with reasons for exclusion will be listed. Inclusion and exclusion criteria failures will be listed. Study completion/withdrawal data will be listed.

#### 6.6 Protocol Deviations

A Protocol Deviation Criteria (PDC) form has been utilised to define the following for any expected protocol deviations:

- Protocol deviation source (either obtained from the database, programmatically and/or via clinical review),
- Protocol deviation category,
- Classification (major/minor),
- Resulting inclusion or exclusion from analysis sets.

This form has been reviewed and approved by Vicore Pharma AB prior to commencing collection of protocol deviations.

All protocol deviations will be assessed and documented on a case-by-case basis. A review of all captured deviations will be completed by Vicore Pharma AB prior to database lock and major deviations considered to compromise the efficacy results will lead to the relevant subject being excluded from the PPAS.

Details of all protocol deviations (date, deviation category, specific details, action taken and classification of major or minor) will be listed. The listing will also include whether a subject was excluded from the PPAS.

#### 6.7 Background and Demographic Characteristics

##### 6.7.1 Demography

Demographic characteristics (age, sex, ethnic origin and race), body measurements (height, weight and body mass index (BMI)) collected at Screening will be summarised. Age (≤median age at baseline, >median age at baseline), supplemental oxygen use at baseline and CRP value at baseline (≤median value at baseline (mg/l), >median value at baseline (mg/l)) will also be presented to align with the subgroup analyses discussed in Section 6.10.6 of this SAP.

Age will be reported using the values entered into the eCRF.

BMI will be calculated as (weight (kg)/height (m)$^2$) using the measurements provided at Screening.

All subject demographic data will be listed including informed consent date and the protocol version enrolled under.

##### 6.7.2 Medical History

Medical history events will be coded using the latest Medical Dictionary for Regulatory Activities (MedDRA) dictionary version. The version used will be indicated in the data summaries and listings. The number and percentage of subjects will be presented for ongoing conditions and previous conditions separately by system organ class (SOC), and preferred term (PT), where SOC and PT will be presented in decreasing frequency of the total number of subjects with medical history events. All events will be listed.

##### 6.7.3 Hospitalisation

The date of admission to hospital and the date of discharge from hospital will be listed. The duration of hospital stay will be derived in days as date of the discharge - date of admission + 1.

##### 6.7.4 Disease Under Investigation

The date of confirmed coronavirus infection diagnosis will be listed. The time from diagnosis to treatment, calculated in days as date of first study treatment administration – date of diagnosis, will also be listed.

#### 6.8 Prior and Concomitant Medications

Medications will be coded using the latest World Health Organization Drug dictionary (WHO Drug) version. The version used will be indicated in the data summaries and listings.

Prior medications are defined as those that started and ended prior to first administration of study treatment. Any medications started on or after first administration of study treatment will be deemed to be concomitant medications. Medications that started prior to first administration of study treatment and are ongoing at the first administration of study treatment will be considered as both prior and concomitant medications. If medication dates are incomplete and it is not clear whether the medication was concomitant, it will be assumed to be concomitant.

The number and percentage of subjects taking prior and concomitant medications will be summarised separately by medication class and standardised medication name, where medication class and standardised medication name will be presented in decreasing frequency of the total number of subjects with medications. Medications which have started prior to first administration of study treatment and are ongoing at the first administration of study treatment will be included in both the summary of prior medications and the summary of the concomitant medications. In summary tables, subjects taking multiple medications in the same medication class or having the same standardised medication recorded multiple times in the study will be counted only once for that specific medication class and standardised medication name.

Medication data will be listed, where concomitant medications will be flagged.

#### 6.9 Administration of Study Treatment and Exposure

For each treatment administration at each visit, the dose administered will be summarised. Dose will be calculated as the number of capsules consumed*50 mg.

The total number of doses of study treatment administered, the treatment duration, actual treatment duration and exposure will be summarised.

Treatment duration will be derived in days as date of the last administration of study treatment - date of first administration of study treatment +1.

Actual treatment duration will be derived as the number of days a subject received study treatment during the treatment period, calculated as treatment duration – number of days of missing administration. Where a subject missed one of the two administrations on a day, they will still be considered to have been treated on that day for this calculation.

Exposure will be calculated as (actual number of doses received/expected number of doses)*100. The expected number of doses will be calculated as treatment duration*2. For example, should a subject complete treatment, the expected number of doses is 14.

Details of the timings of treatment administration, the number of capsules administered and reasons for doses not taken will be listed along with the fasting information confirming that the subject completed the required fasting periods both prior to and following study treatment administration. The treatment duration, actual treatment duration and exposure information will be also listed.

#### 6.10 Efficacy Evaluation

Analysis of the primary endpoint will be performed on the FAS and repeated on the PPAS.

##### 6.10.1 Primary Endpoint

The primary endpoint is the change in CRP from baseline to the mean of the last two non-missing scheduled assessments occurring during the seven day treatment period (Visits 2 to 8 and/or withdrawal). For subjects who do not complete treatment, the assessments up to and including the next-non missing assessment following the final administration of study treatment will be considered for inclusion in the mean.

The baseline CRP assessment is defined as the Visit 2 value supplied by the central laboratory where this assessment occurs prior to the first administration of study treatment. Should the Visit 2 assessment be taken after first administration of study treatment or be missing, the Visit 1 value will not be used and the baseline CRP assessment will remain missing. This avoids potential differences amongst local laboratories that supply the Visit 1 CRP data following the protocol amendment of 14 August 2020. If necessary, the incorporation of Visit 1 local laboratory CRP values may be investigated post-hoc to support any conclusions of the CSR.

Primary Analysis

The null hypothesis will be that the treatments are equivalent and will be rejected in favour of the alternative hypothesis, that a treatment difference exists, should the probability of the null hypothesis being true be less than 10%.

The change in CRP from baseline to the mean of the last two non-missing scheduled assessments during the treatment period will be analysed using an analysis of covariance (ANCOVA). The model will be fitted with the change in CRP from baseline as the dependent variable and study treatment as a fixed effect. The baseline CRP value will be included as a covariate.

Adjusted Least square means (LS Means) for the treatment groups estimated by the model will be presented, together with the associated standard errors (SE) and 90% CIs. The adjusted LS Mean, SE and 90% CI and two-sided p-value of the treatment difference will also be presented.

Model assumptions will be checked. Where these assumptions are not met, data will be log-transformed and the ANCOVA will be performed on the transformed data.

Imputation

Subjects with a missing baseline assessment or no post-baseline assessment of CRP will not be imputed and will not be included in the analysis.

Post-baseline assessments up to and including the next non-missing assessment (including those at or after the withdrawal assessment) following the final treatment administration will be considered as the treatment period for inclusion in the mean. The mean of their last two non-missing scheduled assessments during the treatment period, or a single CRP value where the subject has only completed one post-baseline CRP assessment, will be carried forward to represent state at the end of trial.

Summaries

Descriptive statistics will summarise the CRP values and change from baseline at each assessment for each study treatment. The geometric mean, SD on the log scale and % coefficient of variation (CV) will also be presented. CRP values including the change from baseline will be listed.

Sensitivity Analysis

A sensitivity analysis will be conducted using the mean of the last two non-missing post-baseline values during the treatment and follow-up periods (up to and including Visit 9). If there is only one post-baseline value then this value will be used. This approach is expected to be conservative as it includes a time period when patients in the active treatment arm are potentially untreated.

The analysis will be a repeat of the ANCOVA described above.

Further sensitivity analyses may be considered post-hoc.

##### 6.10.2 Secondary Endpoints

All secondary efficacy endpoints will be analyzed and summarized using the FAS as the primary analysis set.

###### 6.10.2.1 Change from Baseline in Body Temperature and Laboratory Values

Analysis

The change from baseline in body temperature, cytokines (IL-6, IL-10 and TNF), CA125 and ferritin will be analysed using the model outlined for the primary endpoint in Section 6.10.1 of this SAP.

In each case, the change in each parameter from baseline to the mean of the last two non-missing scheduled assessments during the treatment period will be analysed using an ANCOVA. For subjects who do not complete treatment, the assessments up to and including the assessment on the day of last study treatment administration will be considered for inclusion in the mean. The model will be fitted with the change from baseline as the dependent variable and study treatment as a fixed effect. The baseline value will be included as a covariate. Model assumptions will be checked and data transformations will be applied as described in Section 6.10.1.

These analyses will be reported as described for the primary endpoint in Section 6.10.1.

Imputation

Data will be imputed for each parameter as detailed in Section 6.10.1 for the primary endpoint. For subjects who have no post-baseline assessments, no imputation will be applied and they will be excluded from the analysis.

Summaries

Descriptive statistics for observed and change from baseline values of each parameter will be provided at each assessment for each study treatment as outlined for the primary endpoint. Listings of each parameter will be provided including the change from baseline.

###### 6.10.2.2 Number of Subjects Not in Need of Oxygen Supply

A subject will be deemed to not need oxygen if they did not receive supplemental oxygen on their last day of study treatment administration, as recorded on the Supplemental Oxygen eCRF page.

Analysis

The proportion of subjects not in need of oxygen supply at their last study treatment administration visit will be analysed using a logistic regression model. The model will be fitted with oxygen required (Yes/No) as the dependent variable and study treatment as a fixed effect. The number and percentage of subjects not requiring oxygen, together with the proportion of subjects not requiring oxygen and the Clopper-Pearson 90% CI for the proportion, will be presented for each dose group. The odds ratio, associated 90% CI and two-sided p-value will also be presented.

Imputation

For subjects who have withdrawn from treatment due to a need for mechanical ventilation or due to death, or who require mechanical ventilation or die on their last day of study treatment administration, they will be assumed to require oxygen regardless of the oxygen requirement entered into the Supplemental Oxygen eCRF page. For subjects who withdraw from treatment for any other reason, their oxygen status on the last day of study treatment administration according to the Supplemental Oxygen eCRF page will be used. For subjects who have missing data not due to withdrawal from treatment, their last known oxygen status prior to the missing assessment will be carried forward. For subjects who have no post-baseline assessments, no imputation will be applied and they will be excluded from the analysis.

Sensitivity Analysis

A sensitivity analysis will be performed including subjects who have no post-baseline assessments. These subjects will be assumed to require oxygen if they received C21 and assumed not to require oxygen if they received placebo.

The logistic regression model described above will be repeated.

###### 6.10.2.3 Number of Subjects Not in Need of Mechanical Invasive or Non-Invasive Ventilation

The decision for a subject's need for mechanical ventilation (invasive or non-invasive) during the treatment period will determine their response for use in this analysis, irrespective of whether they subsequently received ventilation. A subject will be considered to need mechanical ventilation during the treatment period if they are withdrawn from treatment with the reason for withdrawal recorded as need for mechanical invasive or non-invasive ventilation or death. Any mechanical invasive or non-invasive ventilation or death recorded in the time between last treatment administration + 12 hours will also be considered an event. The treatment period for each subject is from the date and time of their first study treatment administration to the date and time of their last study treatment administration + 12 hours.

Analysis

The proportion of subjects not in need of mechanical ventilation (either invasive or non-invasive) at any point during the study treatment period will be analysed using a logistic regression model. The model will be fitted with ventilation required (Yes/No) as the dependent variable and study treatment as a fixed effect. The number and percentage of subjects not requiring ventilation, together with the proportion of subjects not requiring ventilation and the Clopper-Pearson 90% CI for the proportion, will be presented for each dose group. The odds ratio, associated 90% CI and two-sided p-value will also be presented.

Imputation

Where subjects have been withdrawn from treatment for a reason other than need for mechanical invasive or non-invasive ventilation or death, and have not recorded requirement for mechanical invasive or non-invasive ventilation or death during the time from last treatment administration + 12 hours, they will be considered not in need of mechanical ventilation during the treatment period. All subjects who commence treatment will have an End of Treatment eCRF page completed indicating whether or not the subject was withdrawn due to need for mechanical ventilation or death. Therefore, no further imputation is required.

Summaries

The ventilation requirements, including the date and time the decision was made, the date and time the ventilation started and the date and time the ventilation stopped or if the ventilation was ongoing will be listed.

Sensitivity Analysis

A sensitivity analysis will be performed including any additional information gathered between the last study treatment administration and Visit 9 (follow-up period), therefore this analysis will include all data collected from the first treatment administration to the date of study completion or withdrawal. The need for mechanical ventilation is not captured during the follow-up period however the start time of any mechanical ventilation is captured. Therefore, the sensitivity analysis will consider a subject to need mechanical ventilation during the study if any of the following are recorded from the date and time of first study treatment administration to the date of study completion/withdrawal:

- withdrawal from treatment with the reason for withdrawal recorded as need for mechanical invasive or non-invasive ventilation,
- any entry on the Mechanical Ventilation eCRF page indicating that ventilation was required,
- death as recorded on the adverse event or withdrawal from study pages.

The logistic regression model described above will be repeated.

###### 6.10.2.4 Time to Need of Mechanical Invasive or Non-Invasive Ventilation

Time to need of mechanical ventilation (invasive or non-invasive) for each subject is calculated in hours as (Date and time of decision for need of ventilation) – (Date and time of first administration of study treatment). The treatment period for each subject is from the date and time of their first study treatment administration to the date and time of their last study treatment administration + 12 hours. For subjects who withdraw from treatment with the reason for withdrawal as need for mechanical invasive or non-invasive ventilation, the date and time of decision for need of ventilation is taken from the date and time at which the subject withdrew from treatment (as recorded on the End of Treatment page of the eCRF). For subjects who do not withdraw from treatment due to need for mechanical invasive or non-invasive ventilation but who have an entry on the Mechanical Ventilation eCRF page indicating that ventilation was required during the treatment period, the start date and time from this page will be used to calculate time to need of ventilation. For subjects who do not record a need for ventilation but die during the treatment period, the date and time of death as recorded on the Adverse Event page will be used to calculate the time to need of ventilation.

For subjects not requiring ventilation during their treatment period and who do not die during the treatment period, the time to need of mechanical ventilation will be right censored at the date and time of treatment completion + 12 hours. For subjects who withdraw from treatment for reasons other than need for ventilation or death, the time to need of ventilation will be right censored at the date and time of withdrawal from treatment + 12 hours. The date and time of last treatment administration + 12 hours is a censoring event, should a subject be assessed as having need for ventilation after this time it will be disregarded in the analysis.

Analysis

A log-rank test will be performed to compare the time to need of ventilation for each of the study treatments.

Imputation

Dates will be imputed as described above.

Summaries

Descriptive statistics will be presented for the time to need of ventilation by study treatment. The number and percentage of subjects that are censored will be summarised, along with KM estimates for the 25th, 50th and 75th percentiles of time to need of ventilation and corresponding 90% CIs. The time to need of ventilation will also be presented graphically using a KM plot.

Sensitivity Analysis

The analysis will be repeated to include subjects who record the need for mechanical ventilation or death between the last study treatment administration and Visit 9 (follow-up period), therefore this analysis will include all data collected from the first treatment administration to the date of study completion or withdrawal_to further support the results of the study. For subjects who do not record the need for mechanical ventilation or death during the treatment period, but do record one or both of these during the follow-up period, the earliest of the following will be used to calculate the time to need of ventilation: start time and date as entered on the mechanical ventilation eCRF page or time and date of death as entered on the adverse events page. For subjects who do not record a need for ventilation and who do not die at any point following first study treatment administration, the time to need of ventilation will be right censored at the date of withdrawal from study or study completion.

It should be noted that the decision time is used for subjects who record withdrawal due to mechanical ventilation but the start time of mechanical ventilation will be used for subjects who do not withdraw due to mechanical ventilation but do record an entry on the mechanical ventilation eCRF page.

###### 6.10.2.5 Time on Oxygen Supply

The time on oxygen supply is defined as the number of calendar days on which a subject was recorded as receiving oxygen on the Supplemental Oxygen eCRF page from Visit 2 to Visit 8. For subjects who do not complete treatment, the assessments up to and including the day of the last study treatment administration will be considered and remaining assessments will be imputed as described below. Therefore, a discrete number of responses are available from 0 days to 7 days. Subjects are deemed to have received oxygen where it has been administered at any time for any duration on the date of each visit.

Analysis

A comparison between the two study treatments will be performed using a Wilcoxon rank sum test. A two-sided p-value of the treatment difference will be presented.

Imputations

The treatment period for each subject is defined as from the date of first study treatment administration to the date of last study treatment administration. Where subjects have been withdrawn from treatment with the reason for withdrawal recorded as need for mechanical invasive or non-invasive ventilation, or they record need for ventilation (per the mechanical ventilation eCRF page) on the day of last study treatment administration, they will be assumed to require oxygen for the day of withdrawal / last study treatment administration and all subsequent visits up to and including Visit 8 regardless of the oxygen requirement information entered into the Supplemental Oxygen eCRF page. Similarly if a subject withdraws due to death or dies on the day of last study treatment administration they will be assumed to require oxygen for the day of death and all subsequent visits up to and including Visit 8 regardless of the oxygen requirement entered into the Supplemental Oxygen eCRF page. Otherwise, if a subject has missing assessments for any other reason, their last known oxygen status prior to the missing assessment will be carried forward for the analysis. For subjects who have no post-baseline assessments, no imputation will be applied and they will be excluded from the analysis.

Summaries

The number of days supplemental oxygen is required during screening, the treatment period and the follow-up period will be summarised categorically by treatment. The oxygen requirement for each day will be listed with the imputed oxygen status determined for the end of the treatment period.

##### 6.10.3 Exploratory Endpoints

Biomarker data will be exploratory and the analyses data driven. Analyses and presentations of biomarker results will not form part of this SAP.

Biomarker sample collection details will be listed.

##### 6.10.4 Multiplicity

All secondary endpoints and the supportive analyses will be considered as descriptive evidence of efficacy and will be analysed without any procedures to account for multiple comparisons.

##### 6.10.5 Country Effect

The analyses described in Sections 6.10.1 and 6.10.2 will include country in the model as appropriate, where greater than 10 subjects are recruited per country. Summary tables will also be presented by country and overall should greater than 10 subjects be recruited per country.

##### 6.10.6 Subgroup Analyses

The primary efficacy endpoint will be investigated using demographic and baseline value-defined subgroups as follows:

- Age (≤median age at baseline, >median age at baseline).
- Sex (Males, Females).
- Supplemental oxygen use at baseline (Yes, No).
- CRP value at baseline (≤median value at baseline (mg/l), >median value at baseline (mg/l)).

Each subgroup (age, sex, supplemental oxygen use and CRP) will be examined separately.

A summary table will be presented for each category of each subgroup for the primary endpoint. Analysis will be performed as described in the appropriate section for each category of the subgroup separately. Summaries and analyses will only be presented for a subgroup providing greater than 10 subjects are available for each treatment group and each category of the respective subgroup. All summaries and analyses will be presented on the FAS only.

Secondary endpoints may be explored using these subgroups and further subgroups may be also explored for all endpoints post-hoc where deemed medically appropriate following review of the data.

#### 6.11 Safety Evaluation

A secondary endpoint is the incidence of AEs. Details regarding the evaluation of these data are given in the following section.

##### 6.11.1 Adverse Events

Adverse events will be coded using the latest MedDRA dictionary version. The version used will be indicated in the data summaries and listings.

A treatment-emergent adverse event (TEAE) is defined as an AE that started on or after the start of the administration study treatment. If adverse event dates are incomplete and it is not clear whether the adverse event was treatment-emergent, it will be assumed to be treatment-emergent.

The relationship with study treatment will be entered in to the eCRF (related/not related). If the TEAE has a missing relationship it is assumed to be related to the study treatment for analysis purposes.

A summary table will present the following:

- TEAEs (events and subjects).
- Serious TEAEs (events and subjects).
- Serious study treatment-related TEAEs (events and subjects).
- TEAEs by severity (mild/moderate/severe) (events and subjects).
- TEAEs by relationship to study treatment (events and subjects).
- TEAEs leading to withdrawal from study (subjects only).
- TEAEs leading to discontinuation of study treatment (subjects only).
- Study treatment-related TEAEs leading to discontinuation of study treatment (subjects only).
- TEAEs leading to death (subjects only).

In the above summaries, if a subject experienced more than one TEAE, the subject will be counted once using the most related event for the “by relationship to study treatment” and “related to study treatment” summaries and at the worst severity for the “by severity” summary.

The following tables will be presented:

- TEAEs by SOC and PT.
- TEAEs by SOC, PT and severity.
- TEAEs by SOC, PT and relationship to study treatment.
- TEAEs leading to treatment withdrawal by SOC and PT.
- Serious TEAEs by SOC and PT.

For all of the above, SOC and PT will be presented in decreasing frequency of the total number of subjects with TEAEs.

Further details of the above five tables are given below:

1. If a subject experienced more than one TEAE, the subject will be counted once for each SOC and once for each PT.
2. If a subject experienced more than one TEAE, the subject will be counted once for each SOC and once for each PT at the worst severity.
3. If a subject experienced more than one TEAE, the subject will be counted once for each SOC and once for each PT using the most related event.

Adverse event data will be listed in full and this will also include a treatment emergent flag, the time of onset and cessation of event relative to first dosing of study treatment and duration of AE.

The listing will be repeated for serious AEs and AEs leading to treatment withdrawal.

##### 6.11.2 Clinical Laboratory Evaluation

Observed values and change from baseline in haematology, biochemistry and urinalysis assessments will be summarised over time. If the test results are reported in categorical format, the results will be summarised by subject counts and percentage for each category.

Each haematology, biochemistry and urinalysis parameter will be classed as low, normal, high, missing based on the reference ranges. Shift tables in relation to the normal range from baseline over time will be presented.

Haematology, biochemistry and urinalysis data will be listed separately including change from baseline, reference ranges flagging all out of range values and their clinical significance.

Listings of clinically significant haematology and biochemistry laboratory measurements recorded throughout the study will be provided.

##### 6.11.3 Viral Serology

Viral serology values will be listed.

##### 6.11.4 Vital Signs

Vital sign observed values and change from baseline by parameter (unit) will be summarised over time.

All vital sign data will be listed including change from baseline.

##### 6.11.5 Electrocardiography

Details of the electrocardiogram (ECG) interpretation at Screening will be listed.

##### 6.11.6 Physical Examination

Abnormal physical examination findings will be included within medical history if identified prior to first administration of study treatment, or within adverse events if identified after first administration of study treatment. They will not be summarised independently. Details of timings of physical examinations will be listed.

###### 6.11.7 Pregnancy Test

Pregnancy test details, including the date and method of sampling, will be listed.

###### 6.11.8 Visit Dates

Dates of each visit for each subject will be listed.

##### 6.12 Changes from the Protocol Planned Analysis

- The SAS definition has been amended to clarify that subjects do not need to be randomised to be included. A subject will be included if they have received study treatment, irrespective of their randomisation.
- The secondary endpoint of time on oxygen supply has been amended to include all subjects and not just those who do not require invasive or non-invasive mechanical ventilation.
- Abnormal physical examinations will not be summarised. This data forms part of medical history or adverse events and will be summarised within these data.

### Document History

SignNow E-Signature Audit Log

All dates expressed in MM/DD/YYYY (US)

**Document name:** VP-C21-006 OCH20005 SAP Final 1.0
**Document created:** 09/10/2020 13:21:34
**Document pages:** 20
**Document ID:** 9c06cb3f2cd942b3b52c7629656ed9f207054180
**Document Sent:**
**Document Status:** Signed
09/10/2020 13:39:20UTC

**Sender:**
**Signers:**
**CC:**

| Client | Event | By | Server Time | Client Time | IP Address |
|---|---|---|---|---|---|
| SignNow Web Application | Uploaded Document | | 09/10/2020 13:21:34 pm UTC | 09/10/2020 13:21:30 pm UTC | 62.232.1.93 |
| SignNow Web Application | Document Saved | | 09/10/2020 13:21:35 pm UTC | 09/10/2020 13:21:30 pm UTC | 62.232.1.93 |
| SignNow Web Application | Viewed the Document | | 09/10/2020 13:22:46 pm UTC | 09/10/2020 13:22:46 pm UTC | 62.232.1.93 |
| SignNow Web Application | Signer Authentication Success | | 09/10/2020 13:22:52 pm UTC | 09/10/2020 13:22:51 pm UTC | 62.232.1.93 |
| SignNow Web Application | Signed the Document, Signature ID: 411ef847d07d44998e00 | | 09/10/2020 13:23:09 pm UTC | 09/10/2020 13:23:08 pm UTC | 62.232.1.93 |
| SignNow Web Application | Added a Text | | 09/10/2020 13:23:09 pm UTC | 09/10/2020 13:23:08 pm UTC | 62.232.1.93 |
| SignNow Web Application | Document Saved | | 09/10/2020 13:23:09 pm UTC | 09/10/2020 13:23:08 pm UTC | 62.232.1.93 |
| SignNow Web Application | Signer Authentication Success | | 09/10/2020 13:38:06 pm UTC | | 36.255.158.14 |
| SignNow Web Application | Viewed the Document | | 09/10/2020 13:38:48 pm UTC | 09/10/2020 13:38:48 pm UTC | 36.255.158.14 |
| SignNow Web Application | Signed the Document, Signature ID: 2f9727c9ebde416f8729 | | 09/10/2020 13:39:20 pm UTC | 09/10/2020 13:39:20 pm UTC | 36.255.158.14 |
| SignNow Web Application | Document Saved | | 09/10/2020 13:39:20 pm UTC | 09/10/2020 13:39:20 pm UTC | 36.255.158.14 |
